# High titre neutralizing antibodies in response to SARS-CoV-2 infection require RBD-specific CD4 T cells that include proliferative memory cells

**DOI:** 10.1101/2022.07.22.22277947

**Authors:** Chansavath Phetsouphanh, Weng Hua Khoo, Katherine Jackson, Vera Klemm, Annett Howe, Anupriya Aggarwal, Anouschka Akerman, Vanessa Milogiannakis, Alberto Ospina Stella, Romain Rouet, Peter Schofield, Megan L. Faulks, Hannah Law, Thidarat Danwilai, Mitchell Starr, C. Mee Ling Munier, Daniel Christ, Mandeep Singh, Peter I Croucher, Fabienne Brilot-Turville, Stuart Turville, Tri Giang Phan, Gregory J Dore, David Darley, Philip Cunningham, Gail V Matthews, Anthony D Kelleher, John J Zaunders

## Abstract

Long-term immunity to SARS-CoV-2 infection, including neutralizing antibodies and T cell-mediated immunity, is required in a very large majority of the population in order to reduce ongoing disease burden. We have investigated the association between memory CD4 and CD8 T cells and levels of neutralizing antibodies in convalescent COVID-19 subjects. Higher titres of convalescent neutralizing antibodies were associated with significantly higher levels of RBD-specific CD4 T cells, including specific memory cells that proliferated vigorously in vitro. Conversely, up to half of convalescent individuals had low neutralizing antibody titres together with a lack of receptor binding domain (RBD)- specific memory CD4 T cells. These low antibody subjects had other, non-RBD, spike-specific CD4 T cells, but with more of an inhibitory Foxp3+ and CTLA-4+ cell phenotype, rather than the effector T- bet+, cytotoxic granzymes+ and perforin+ cells seen in high antibody subjects. Single cell transcriptomics of antigen-specific CD4+ T cells from high antibody subjects revealed heterogenous RBD-specific CD4+ T cells that comprised central memory, transitional memory and Tregs, as well as cytotoxic clusters containing diverse TCR repertoires, that were absent in individuals with low antibody levels. However, vaccination in low antibody convalescent individuals led to a slight but significant improvement in RBD-specific memory CD4 T cells and increased neutralizing antibody titres. Our results suggest that targeting CD4 T cell epitopes proximal to and within the RBD- region should be prioritized in booster vaccines.

**One Sentence Summary:** Individuals with low neutralising antibody titres may be at risk of SARS-CoV-2 re-infection due to a failure to generate a high quality CD4 T cell response specific for receptor binding domain (RBD), including memory CD4 T cells that proliferate in vitro in response to RBD, and which are also therefore an important target for vaccine design.

## INTRODUCTION

The question of the protective efficacy of both convalescent and vaccine induced immunity to SARS-CoV-2 is of global significance. This is highlighted by the multiple waves of infections, with rates of population-wide protection against re-infection of only 50-80% protection during 2020, depending on age (*1*), or an estimated 60% protection against symptomatic re-infection with the immuno-evasive Omicron variant (*2*). In order to end the COVID-19 pandemic, a large proportion of the population will need to be immune to the virus (*3*), or at least, if they become infected, to develop an immune response which minimizes virus shedding, symptoms and onward transmission. Early studies concentrated on serum antibody levels in recovered COVID-19 patients, and in particular neutralizing antibodies, since it is believed that the most effective vaccines to viral infections are associated with the generation of neutralizing antibodies and mimic natural infection (*4*). The highest titres and affinities of such neutralizing antibodies are generally dependent on CD4 T follicular helper cell (Tfh) interaction with B cells to generate class switching and affinity maturation by somatic hypermutation within germinal centres, in secondary lymphoid organs (*5*).

A recent study of the immune response to the mRNA COVID-19 vaccine in SARS-CoV-2 naïve and recovered individuals showed a rapidly induced CD4 T cell response when compared to the gradually developing CD8 T cell response (*6*). Similarly, when we studied the immune response to vaccinia virus inoculation, highly activated antigen-specific CD4 T cells were often more expanded than corresponding CD8 T cells in blood early in the response (*7, 8*). Antigen-specific CD4 cytotoxic T lymphocytes (CTL) peaked at day 14, while neutralizing antibodies, and memory CD4 T cells that proliferated in vitro in response to vaccinia antigen appeared later at day 21 (*7, 8*). The early generation of CD127 (IL-7 receptor alpha chain)-expressing, IL-2-producing, proliferative memory CD4 T cells specific for vaccinia virus is most likely crucial to the long-term protection associated with immunity to smallpox (*9*). Such T cell proliferation in response to re-exposure to viral antigens is believed to be critical to allow more rapid response to re-infection, after the immune system had returned to homeostasis following contraction of the initial response to the acute infection (reviewed in (*10*)). Overall, in vitro proliferation of PBMC in response to antigens derived from pathogens is highly correlated with effective immunity (*11*), due to rapid memory recall response.

A number of studies early in the pandemic identified SARS-CoV-2 specific CD4 T cells in COVID-19 patients (*12–16*). However, longer term studies showed a consistent average decline of memory CD4 T cells over time with a decrease until 60 days after the acute phase and maintained over 10 months with a central memory phenotype post 6 month (*17, 18*). Most of these studies have used upregulation of markers of activation on antigen-specific CD4 T cells, similar to the original OX40 assay (*19*). In contrast there is very limited data on proliferative memory SARS-CoV-2-specific CD4 T cells (*17, 20*).

While CD4 T cell responses are important for antiviral humoral immunity, CD8 T cells are still believed to be important in cell-mediated immunity to respiratory viruses (*21*). Studies investigating the properties of SARS-CoV-2 reactive CD8 T cells report a high diversity of funtional SARS-CoV-2 specific CD8 T cells (*22–24*), with cross-reactivity to seasonal coronavirus antigens (*25*). This indicates that CD8 T cells could be important in viral clearance in individuals that lack neutralizing antibodies (*26, 27*), although they often display an exhausted phenotype (*22, 23*).

In the current study we systematically studied SARS-CoV-2-specific proliferation of CD4 and CD8 T cells in recovered COVID-19 patients to better define the extent of their long-term memory cells. Importantly, RBD-specific proliferative memory CD4 T cells were closely associated with levels of neutralizing antibodies in recovered patients and in vaccinees. Furthermore, single cell RNAseq/TCRseq was used to study the function of individual SARS-CoV-2 reactive CD4 T cells from subjects with high antibody titres.

## RESULTS

### Proliferative RBD-specific CD4 T cells correlate with neutralising antibody titres

To assess memory CD4 T cell responses following SARS-CoV-2 infection, we screened 25 ADAPT subjects from the first wave (May-October, 2020; Supplementary Table 1) at 3 months post-infection using recombinant SARS-CoV-2 RBD protein, and using influenza lysate as a control antigen. We utlised our CD25/OX40 (*19*) assay to assess antigen-specificity of CD4 T cells and found overall a 13.5-fold higher response to RBD in ADAPT subjects (median 0.46%) at 3 months compared to unexposed controls (n=13; median 0.034%, *p*<0.0001) (Figure 1A&B). However, 7/27 ADAPT subjects were negative (<0.2%) for a response to RBD (Figure 1B). All unexposed controls and ADAPT subjects had positive flu specific responses (medians 1.51% and 1.62%, respectively).

**Figure 1.**
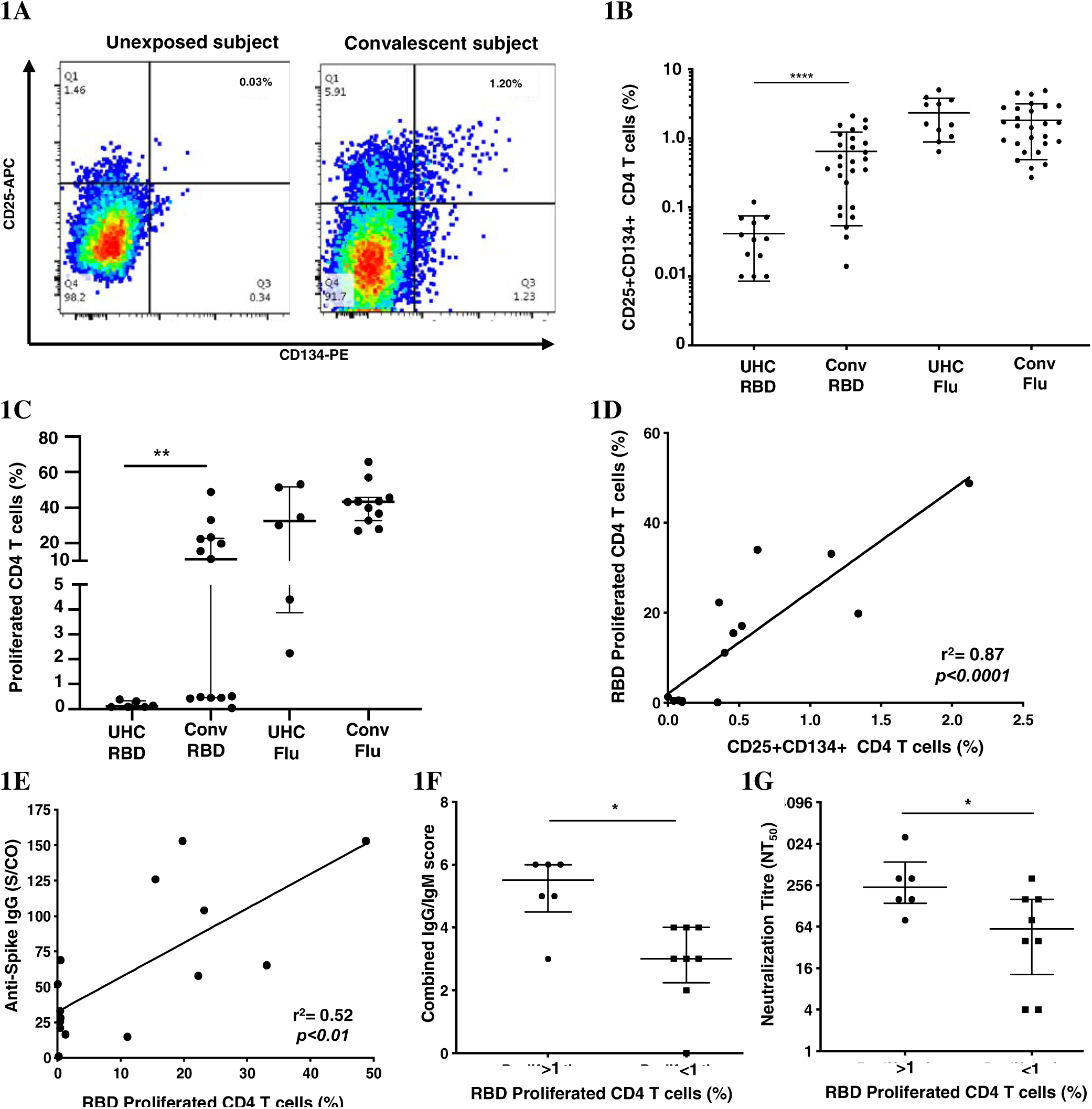
Proliferation of SARS-CoV-2 reactive CD4+ T cells correlate with antibodies. **(A)** Representative dot plots of CD25+CD134+ co-expression on RBD-specific CD4+ T cells. **(B)** Frequency of RBD reactive CD4+ T cells in convalescent subjects (conv) compared to unexposed healthy controls (UHC). **(C)** Percentage of proliferating CD4+ T cells stimulated with RBD or Flu antigen. **(D)** Positive correlation between recall CD25+CD134+ CD4 response and 7 days of proliferation. **(E)** Correlation between spike IgG levels and RBD specific proliferation of CD4+ T cells. **(F)** Convalescent subjects with proliferated CD4+ T cells (>1%) are Ab high *vs* subjects without proliferation (<1%) are Ab low, based on combined IgG/IgM score. **(G)** Subjects with proliferated CD4+ T cells (>1%) have significantly higher neutralizing Ab titres (NT_50_) *vs* subjects without proliferation (<1%). Data shown as medians with interquartile ranges. Two-tailed p values <0.05 were considered significant (*<0.05, **<0.01, ***<0.001, ****<0.0001). Mann-Whitney T tests were used for unpaired groups and Pearson’s rho was used for correlations.

Having seen initial OX40 responses to RBD, we subsequently used the 7-day PBMC proliferation assay with cell trace violet dye (CTV; Supplementary Figure 1) to confirm the presence of memory CD4 T cells. There was no CD4 T cell proliferation response to RBD (< 1% of CD4 T cells) in PBMC from unexposed controls (n=6), while the ADAPT subjects (n=13) had an overall median proliferation of 11.1%, *p*<0.01. Again the results were heterogeneous with 6 of the 13 ADAPT subjects tested having < 1 % CD4 T cell proliferation in response to RBD, similar to the unexposed controls in this assay (Figure 1C). Proliferation responses to Flu lysate, by both control and patient PBMC, were all positive, similar to the OX40 results, and generally larger than the responses to SARS-CoV-2 RBD (Figure 1C). There was a highly significant positive correlation between CD25+ OX40+ CD4 T cell responses to RBD and CD4 T cell proliferation responses to RBD (pearsons’ rho=0.89, *p*<0.0001) (Figure 1D), as we have previously reported for a variety of other recall antigens (*19*).

When we compared the levels of anti-spike IgG, measured in the ADAPT subjects’ 3-month follow-up serum using the diagnostic DiaSorin Liaison assay, the proliferation of RBD-specific CD4 T cells positively correlated with anti-spike IgG levels (rho=0.52, *p*<0.01; Figure 1E). We also used a flow-based assay (*28*) to measure spike (Wuhan-1 D614)-specific IgG and IgM in patients’ serum samples, which gives scores of 0-3 for each antibody isotype (where 3 is highest). When the IgG and IgM scores were combined, the patients with RBD-specific CD4 T cell reponses had significantly higher antibody levels (median score of 5.5) than patients with negative proliferative responses to RBD (median score of 3, *p*<0.05) (Figure 1F).

Finally, live virus neutralization assays were performed with the ADAPT patients’ sera, measured as an end-point titre (*28*). The patients with RBD-specific CD4 T cell reponses had significantly higher neutralization titres (NT_50_, median 256) than patients with negative proliferation results (median 64, *p*<0.05) (Figure 1G). Collectively these data demonstrate a heterogeneous memory immune response observed during natural infection with SARS-CoV-2.

### Antibody High versus Antibody Low subject groups

In order to confirm the association of RBD-specific CD4 T cell responses with higher antibody levels, cryopreserved PBMC and serum samples were studied from an additional 24 ADAPT subjects, separated into two representative groups of 12 subjects each, with known high and low SARS-CoV-2 neutralisation titres, respectively, at month 3 (Supplementary Table 1). The selected antibody high (Ab high) subject group had neutralisation titres NT_50_ ≥ 80, whereas the antibody low (Ab low) group had neutralization titres NT_50_ ≤40 (Figure 2A). Neutralisation titres in the Ab high group gradually decreased over time with a median of 320 at 3-month, 240 at 4-month and 120 at 8-month timepoints (Figure 2B). Furthermore, the breadth of neutralizing antibodies to variants of concern was significantly greater in Ab high group at 3-months (Figure 2C), but also then decreased at 8-months (Figure 2D).

**Figure 2.**
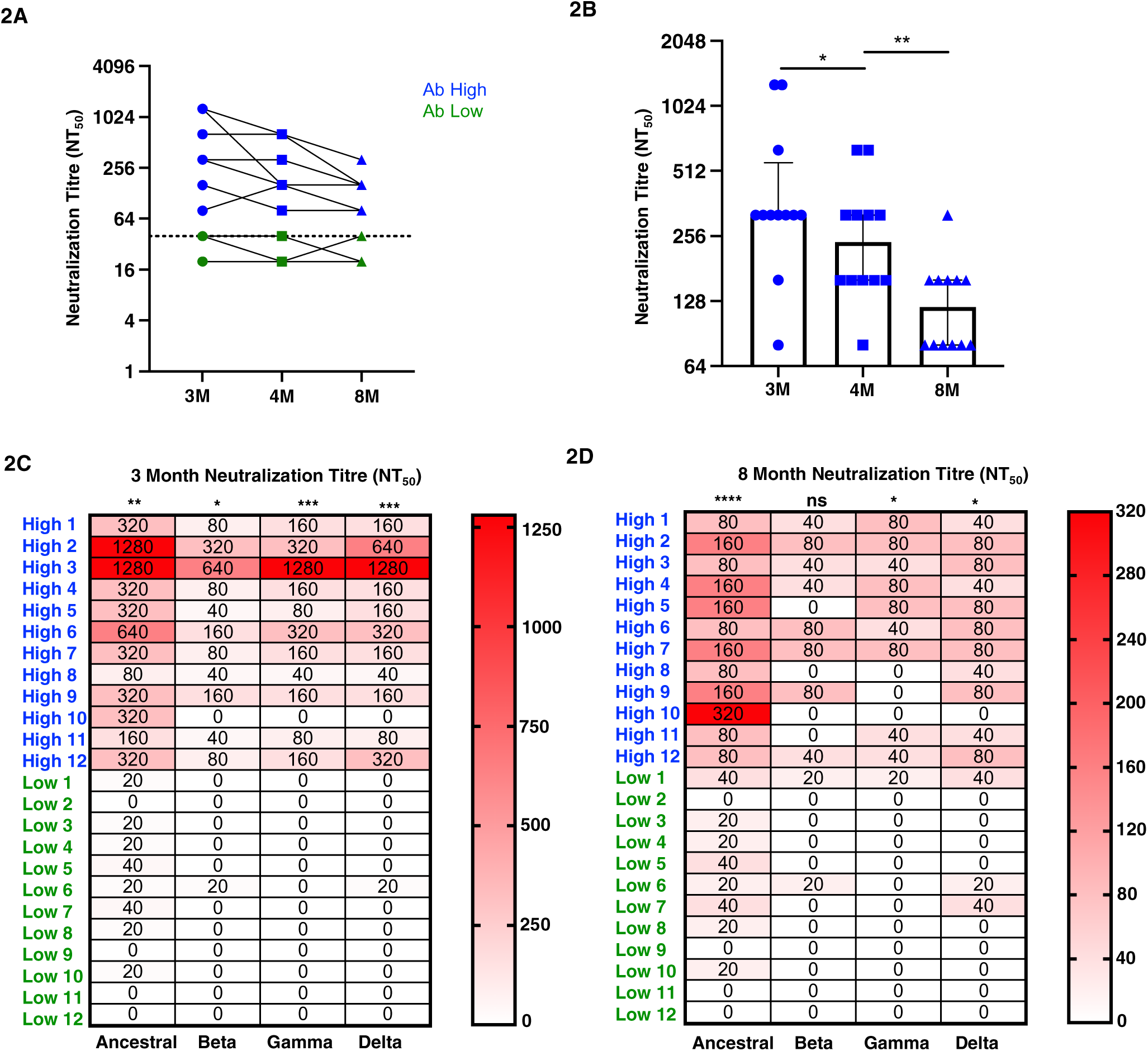
Higher neutralization breadth in Ab high group that decreases over time. **(A)** Representative dot plot showing difference in neutralisation of Ab high (blue) and Ab body low (groups) over time. **(B)** Reduction of neutralizing antibodies over time. 3M (3 months), 4M (4 months), 8M (8 months). **(C)** Increased neutralising breadth of Ab high group compared to antibody low. **(D)** Decreased neutralization tire in Ab high group at 8-months post-infection but remain higher than Ab low. Two-tailed p values <0.05 were considered significant (ns-not significant, *<0.05, **<0.01, ***<0.001, ****<0.0001). Wilcoxon T tests were used for paired groups and Mann-Whitney sued for unpaired. Heatmap values shown as median neutralization titre.

### Reduced RBD-specific CD4 T cell responses in indviduals with low antibody levels

We also widened our analysis of T cell responses to other SARS-CoV-2 antigens, using peptide pools (Supplementary Table 2), that allowed us to adapt our CD25/OX40 CD4 assay to include the detection of antigen-specific CD8 T cells by adding the co-expression of CD69 and CD137 (4-1BB) surface markers (*29*).

RBD-specific CD4 T cells were mostly undetectable in the Ab low group (median 0.12% (3-month) and 0.1% (8-month)), such that the fold difference in RBD-specific CD4 T cell responses between the 2 subject groups was 50-fold at month 3 (*p<*0.001), down to 15-fold at month 8 (*p<*0.01; Figure 3A). There were also significantly higher CD4 T cell responses to the spike peptide pool in the Ab high subject group compared to the Ab low subject group (Figure 3A), although not as marked as for RBD (above), with a 5.3-fold higher spike-specific response at the 3-month time-point (*p<*0.01), which was still 3.9-fold higher at 8 months (*p<*0.01; Figure 3A).

**Figure 3.**
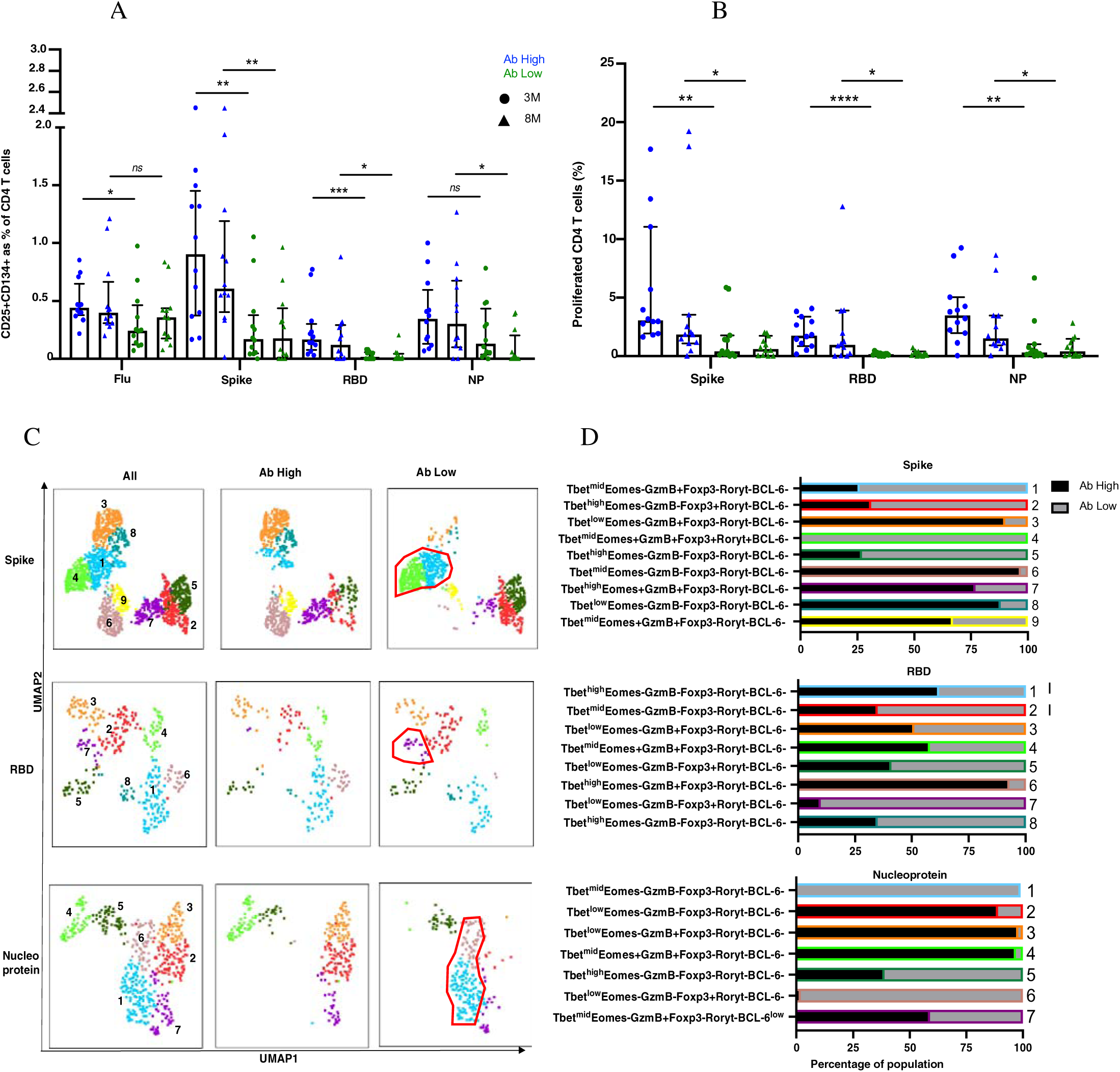
Higher frequencies of RBD-specific CD4+ T cells in Ab high group. **(A)** CD4 responses to influenza, spike, RBD and nucleoprotein (NP) at 3M and 8M post- infection. **(B)** Higher frequency of SARS-CoV-2 proliferative CD4 T cells in Ab high group compared to Ab low at 3M and 8M. (medians with interquartile ranges). **(C)** Representative UMAP of proliferated CD4 T cells: concatenated; ab high; and ab low. Red gates are regulatory-like cells present in Ab low subjects. **(D)** Regulatory-like Foxp3+ cells are higher in Ab low subjects (grey columns) compared with higher effector cells in Ab high subjects (black columns). The colour and number of horizontal bar graphs match with UMAP clusters. Two-tailed p values <0.05 were considered significant (ns-not significant, *<0.05, **<0.01, ***<0.001, ****<0.0001). Mann-Whitney T test used for unpaired samples.

Nucleocapsid protein (NP)-specific CD4 T cell responses were not significantly different between the 2 groups at 3-months, but there was a 15-fold higher frequency in the Ab high subject group compared to the Ab low subject group at 8-months (*p<*0.01). No difference between the 2 groups was observed in the proportion of antigen-specific CD4 T cells that had a CD39+ Treg phenotype (*30*) (Supplementary Figure 2).

In addition, we also measured proliferation of CD4 T cells following peptide stimulation and found that there was significantly higher proliferating SARS-CoV-2-reactive CD4 T cells in the Ab high group at both 3- and 8-month timepoints for all 3 peptide pools (Figure 3B). Intracellular phenotyping and dimenisonal reduction utilising UMAP algorithm of spike-specific proliferated CD4 T cells from the Ab high group showed 3 relatively large clusters of Granzyme B+ cells (clusters #3, #7 and #9; Figure 3C&D, top row) and relatively few regulatory-like cells expressing Foxp3 (clusters #2 and #4; Figure 3C&D, top row). In contrast, proliferated CD4 T cells, in response to the spike peptide pool, from subjects in the Ab low group revealed the greatly increased presence of the 2 clusters of regulatory-like cells expressing Foxp3 (clusters #2 and #4, Figure 3C&D, top row), and greatly reduced Granzyme B+ cells (clusters #3, #7 and #9, Figure 3C&D, top row), compared to the proliferated CD4 T cells from subjects in the Ab high group.

Similarly, in RBD-specific proliferated CD4 T cells, the Ab high group had an increased presence of a Granzyme B+Foxp3- cluster (#6; Figure 3C&D middle rows) and less cells in Foxp3+ clusters (#5 and #7; Figure 3C&D middle rows). The opposite was seen in the much smaller proportions of proliferated cells from the Ab low group (Figure 3C&D middle rows). Nucleoprotein-specific proliferated CD4 T cells also showed a very similar trend, with distinct Foxp3+ versus Granzyme B+ clusters between the Ab high and low groups (Figure 3C&D bottom rows).

In CD8 T cell specific responses, defined by upregulation of CD137 and CD69, no significant difference was observed between the antibody high or low groups, when stimulated with the 3 SARS-CoV-2 peptide pools or influenza (Figure 4A). However, in proliferation responses, the antibody high group had 4.2-fold higher proliferating NP peptide pool-specific CD8 T cells at 3 months (*p<*0.01) and 6.6-fold at 8 months (*p<*0.05), and a trend to higher spike-specific proliferation, compared to the antibody low group (Figure 4B). The much smaller proportions of proliferated CD8 T cells from Ab low subjects had clusters with much higher levels of expression of the inhibitory ligand CTLA-4, compared to proliferated CD8 T cells from Ab high subjects for all 3 peptide pools (Figure 4C&D).

**Figure 4.**
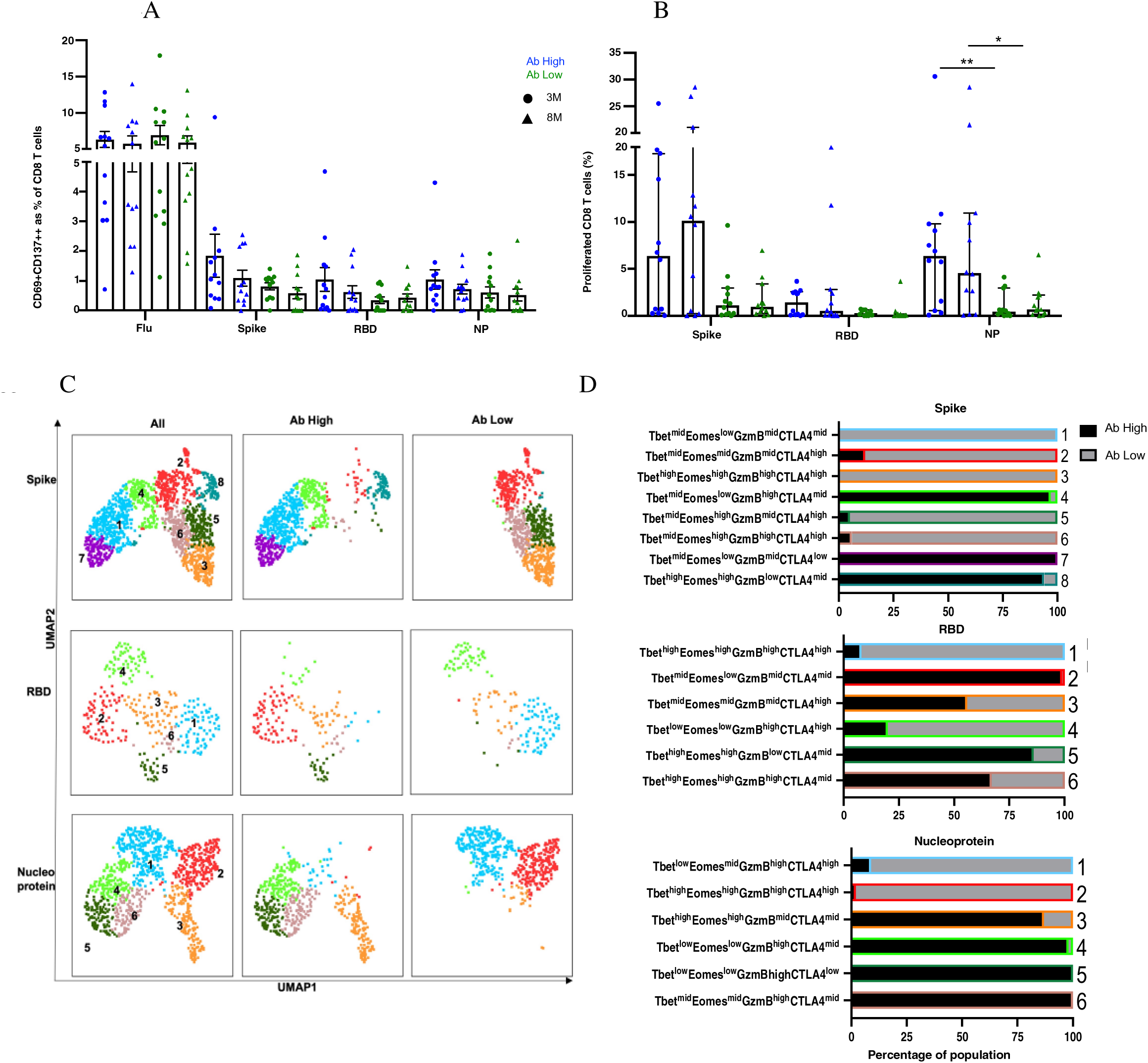
No difference in CD8 responses but presence of inhibitory receptors in Ab low subjects. **(A)** No difference between CD8 in Ab high and Ab low groups. **(B)** Higher frequency of NP proliferative CD8 T cells in Ab high group compared to Ab low. 3M (3 months) and 8M (8 months). Data shown as medians with interquartile ranges. **(C)** Representative UMAP of proliferated CD8 T cells; concatenated, ab high and ab low. **(D)** Presence of an inhibitory phenotype within cells of Ab low subjects. Colour and number of horizontal bar graphs match with UMAP clusters. Black columns (Ab high) and grey columns (Ab low). Two-tailed p values <0.05 were considered significant (*<0.05, **<0.01). Mann-Whitney T test used for unpaired samples.

### B cell plasmablasts, with more CXCR5+ CD4 T cells, and T cell activation are associated with high antibody levels

We used 20-parameter flow cytometry to determine ex vivo phenotypic differences in the B and T cell subsets of Ab high and Ab low groups. Higher frequencies of B cell plasmablasts (CD19+IgD- CD27+CD38+) were found in the Ab high group (3.2-fold, *p*<0.05) at 3 months, but no difference was observed at 8 months (Figure 5A). When CD19+ B cells were divided into memory subsets, there was no difference at either time point for naïve (IgD+CD27-), non-switched memory (IgD+CD27+) or switched memory (IgD-CD27+)(Figure 5B). However, there was higher proportion of double negative (IgD-CD27-) cells in Ab low group at both 3- (1.7-fold, *p*<0.05) and 8-months (1.6-fold, *p*<0.05).

**Figure 5.**
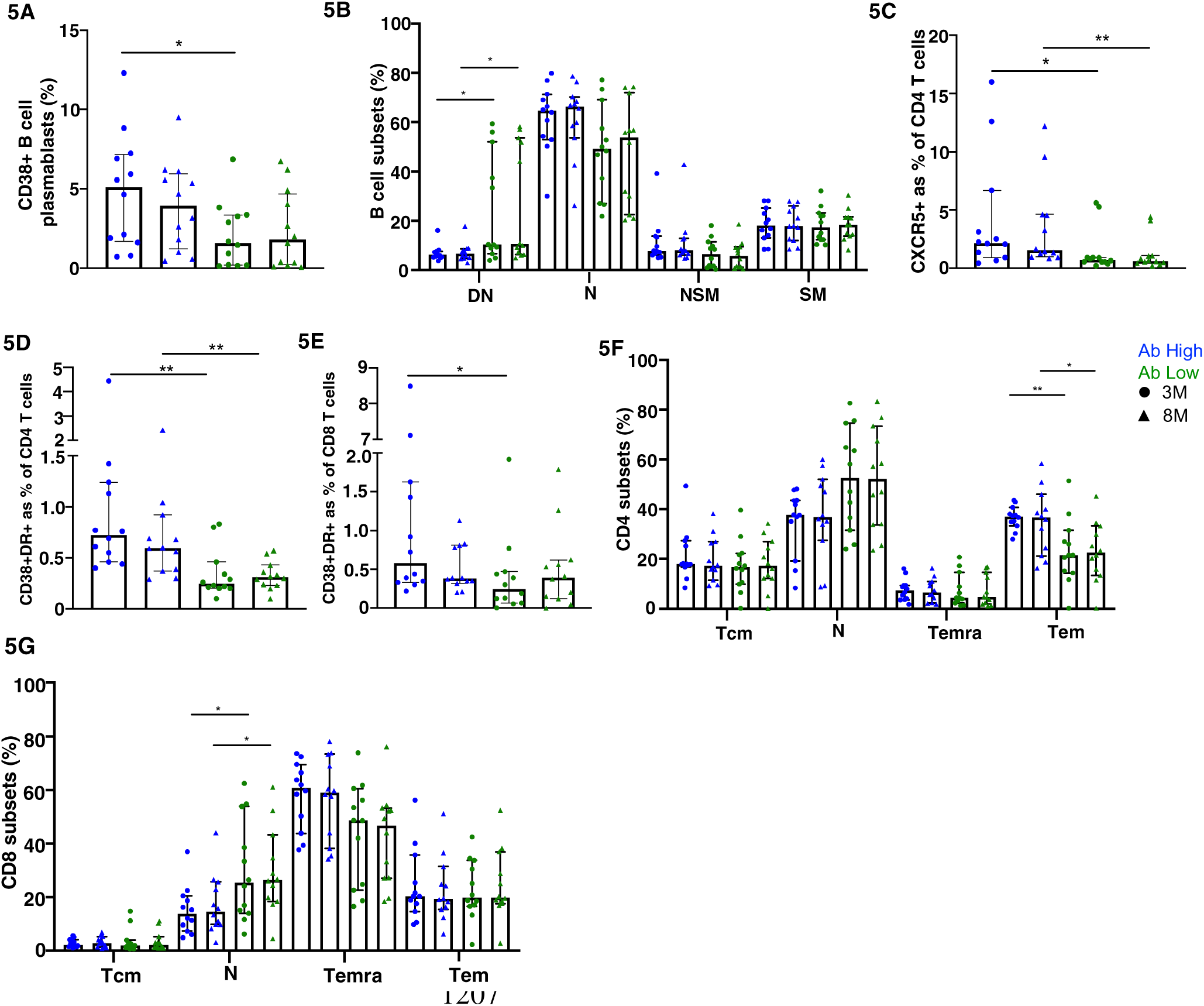
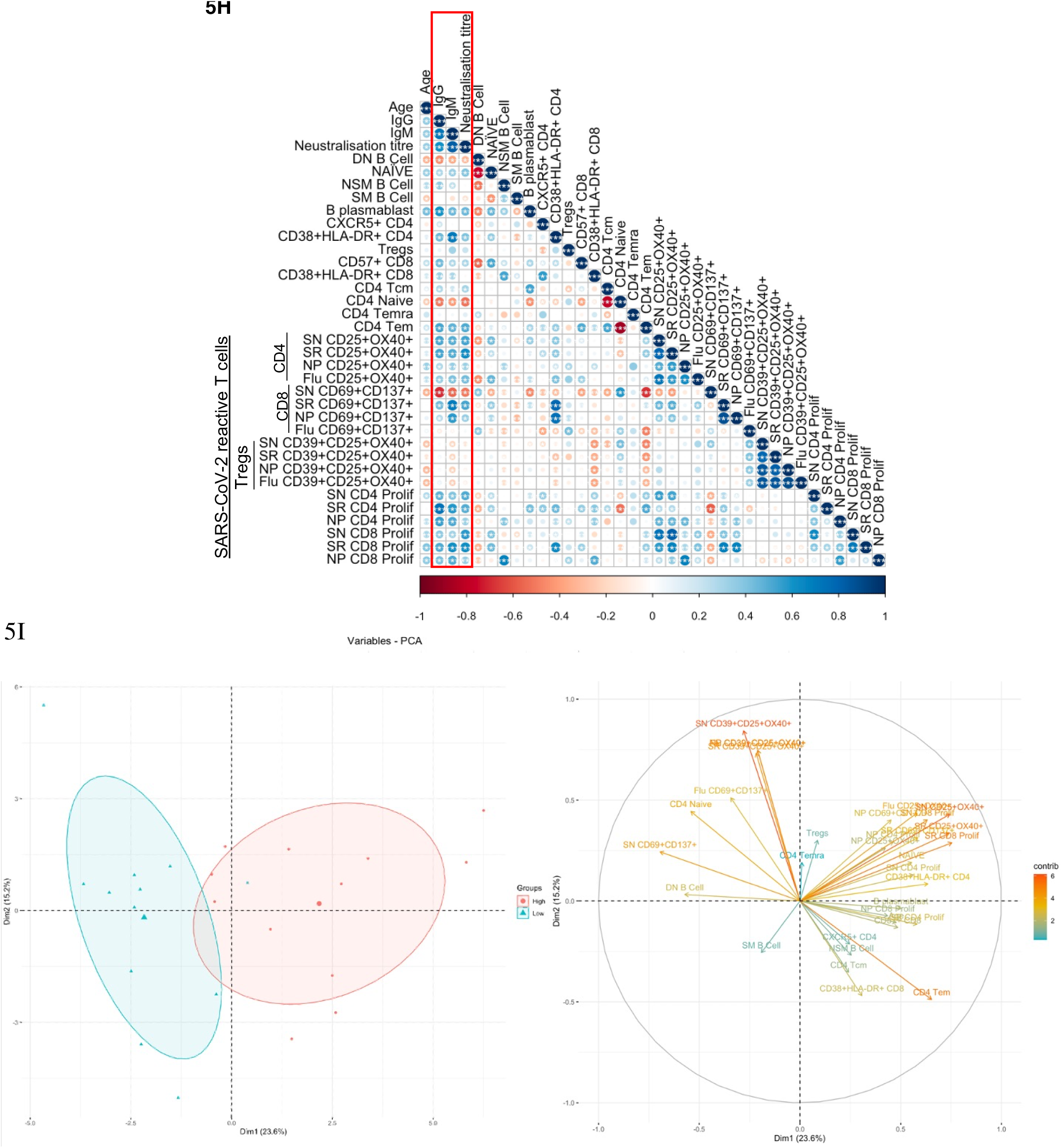
B and T cell parameters correlate with antibody response. **(A)** Higher CD38+ B cell plasmablasts in Ab high group. **(B)** Elevated double negative (DN, IgD-CD127-) B cells in Ab low group. Nailve (N, IgD+CD127-), non-switched memory (NSM, IgD+CD127+), switched memory (SM, IgD-CD127+) B cells. Higher CXCR5 expression on CD4s **(C)**, activation (CD38+HLA-DR+) on CD4 **(D)** and CD8 **(E),** and CD4 effector memory cells **(F)** in Ab High compared to Ab low group. Data shown as medians with interquartile ranges. **(G)** Higher nailve CD8+ T cells in Ab low group at both timepoints. SN (spike), SR (RBD), NP (nucleoprotein). Two-tailed p values <0.05 were considered significant (*<0.05, **<0.01, ***<0.001. Mann-Whitney T tests were used for unpaired groups. **(H)** B cell plasmablast, CXCR5+ CD4 T cells, activated CD4 and CD8 T cells, proliferative Spike and RBD- specific CD4 positively correlate with humoral response at 3 months post-infection (red box). **(I)** PCA clustering of Ab high and Ab low groups based on T and B cell parameters. Pearson’s rho was used for correlations and adjusted *p* values shown with Benjamini-Hochberg correction used for multiple comparisons. Two-tailed p values <0.05 were considered significant (*<0.05, **<0.01, ***<0.001.

When surface markers of CD4 T cells were analysed there was significantly increased expression of CXCR5 in the Ab high group at both 3- and 8-month timepoints (2.9-fold, *p*<0.05 and 2.5-fold, *p*<0.01, respectively) and co-expressed activation markers HLA-DR+CD38+ (3-fold, *p*<0.01 and 1.9- fold, *p*<0.01, respectively), compared to the Ab low group (Figure 5C&D). Activated CD8 T cells were also increased at 3-months in Ab high subjects (2.3-fold, *p*<0.05) but not at 8-months (Figure 5E).

Within canonical CD4 T cell subsets, a higher frequency of effector memory cells (Tem; CD45RA- CCR7-) was evident in the Ab high group (1.7-fold, *p*<0.05 (3-months) and 1.6-fold, *p*<0.05 (8- months), compared to the Ab low group, but no difference was seen in naïve (CD45RA+CCR7+), central memory (Tcm; CD45RA-CCR7+) or memory revertant cells (Temra; CD45RA+CCR7- ; Figure 5F). However, naïve (CD45RA+CCR7+) CD8 T cells were significantly higher in the Ab low group (1.8-fold, *p*<0.05), compared to the Ab high group, at both timepoints (Figure 5G).

To ascertain the association between the results of the T cell function assays and antibody levels, Spearman’s correlation was performed utilising Benjamini-Hochberg method to correct for multiple comparisons. Positive correlations of antibody levels and neutralization titres were observed with: B cell plasmablasts; CXCR5+ CD4 T cells; CD4 and CD8 T cell activation; and spike- and RBD-specific CD4 T cell recall and proliferative responses. In contrast, DN B cells, naïve CD4 and spike-specific CD8 recall responses were all negatively correlated with antibodies (Figure 5H). These same parameters were able to separate Ab high and low groups using PCA (Figure 5I).

### Single-cell RNAseq revealed heterogeneity and TCR diversity in SARS-CoV-2 reactive CD4 T cells

Our previous observations showed that CD4 T cell recall responses to SARS-CoV-2 positively correlated with greater humoral responses, in particular RBD-specific responses in the Ab high subject group. To better understand these antigen-specific CD4 T cells, especially those specific for RBD, we sorted CD25+CD134+ CD4 T cells from 4 representative subjects from the Ab high group, following stimulation with peptide pools at 44hrs. Unstimulated purified memory CD4 T cells were used as the comparison population (Supplementary Figure 3A&B). Single-cell RNAseq was performed on a total of 14,053 cells comprising: (i) 9,039 ex vivo (unstimulated) memory cells; (ii) 2,026 NP-specific cells; (iii) 1,989 spike-specific cells; and (iv) 999 RBD-specific cells. Analysis of 10x transcriptomics was coupled with TCRα/β sequencing. We found heterogeneous subsets of CD4 T cells that were reactive to spike, RBD and NP; comprising transitional memory, central memory, Treg and cytotoxic subsets (Figure 6 A&B). Enrichment of activated GITR+ Tregs (cluster 3) was particularly evident in the stimulated conditions (48.15%, 62.34% and 54.94% for NP- Spike- RBD-specific cells, respectively), compared to 3.08% for ex vivo memory cells (Figure 6C).

**Figure 6.**
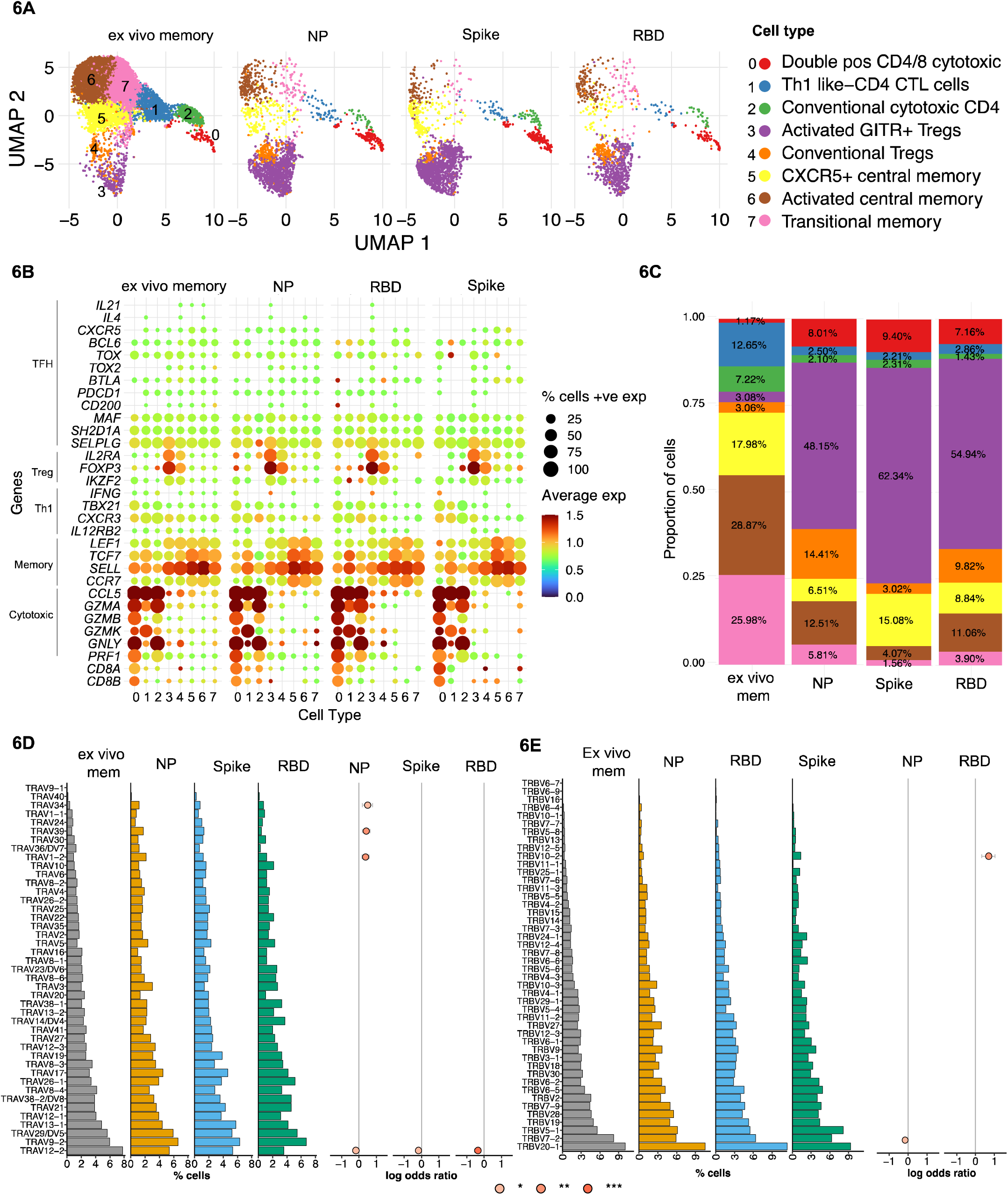
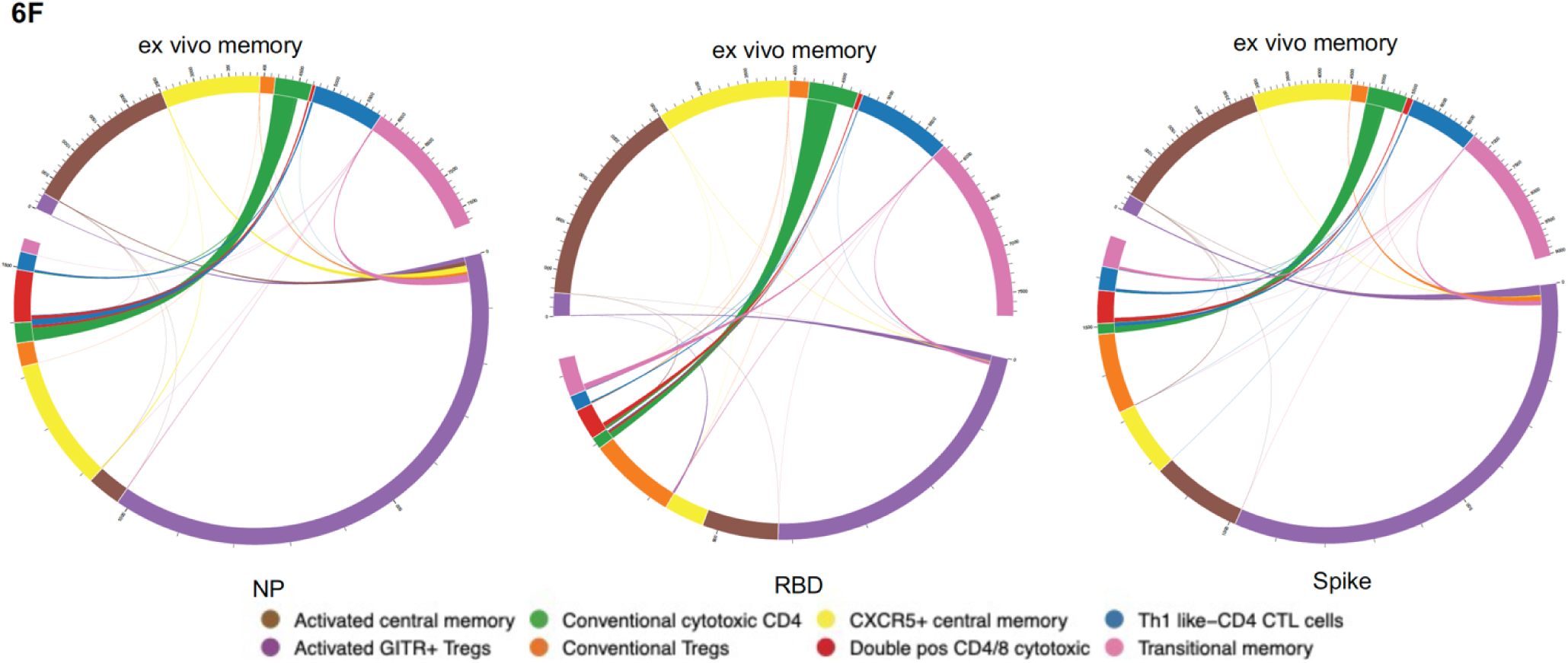
Single-cell RNAseq reveals heterogenous subsets and diverse TCR usage within SARS-CoV-2 reactive CD4+ T cells. **(A)** UMAP showing 14,053 CD4+ T cells from CD25+CD134+ sorted cells from Ab high subjects split into 4 conditions: ex vivo memory; nucleoprotein (NP); Spike; and RBD. **(B**) Dot plot showing average expression of lineage genes and percentage of cellular expression within each cluster. **(C)** Enrichment of Treg populations in SARS-CoV-2-specific CD4 in addition to central/transition memory and cytotoxic subsets. **(D)** Diversity of TCR alpha and **(E)** TCR beta chain in all 4 conditions. Few select TCRa/b enriched in stimulated conditions compared to ex vivo memory expressed as log odds ratio with adjusted *p* values including Bonferroni correction. **(F)** Circos plots showing little transition of cell types between ex vivo memory and stimulated conditions.

Analysis of TCR annotation was achieved on 11,824 single cells (84.14%). Diverse TCR responses resulted from all 3 stimulation conditions with similar Shannon Entropy score for the stimulation conditions compared to the unstimulated ex vivo (Supplementary Figure 4A) and diverse TCRα/β pairings for all four conditions (Supplementary Figure 4B). To examine if TCRα/β V gene was altered in the stimulated conditions compared to ex vivo memory, log odds-ratio analysis was performed with Bonferroni’s correction. No difference was observed in the majority of TCRα/β V genes between ex vivo memory and stimulated conditions. However, there was increased usage of TCRα TRAV1- 2/34/39 chains evident in NP-specific CD4 T cells compared to ex vivo memory (Figure 6D), while TRBV10-2 was the only TCRβ chain enriched in RBD-specific CD4 T cells (Figure 6E). With the exception of CD4 CTLs, where approximately half of the ex vivo unstimulated clones were re-sampled post-stimulation, generally antigen-specific clonotypes within the ex vivo unstimulated memory cell pool comprised only a tiny fraction of the antigen-specific clonotypes sampled following 2 days of antigen stimulation (Figure 6F). Clonotypes shared between unstimulated ex vivo CD4 memory and antigen specific pools rarely displayed altered cellular phenotypes following stimulation (Figure 6F).

### Proliferative CD4 T cell clonotypes are enriched within cytotoxic and Treg subsets

To examine the clonal diversity of proliferative CD4 T cell subsets and confirm the single cell RNAseq clonotype data, we performed bulk TCR sequencing on T cells proliferating in response to NP, spike and RBD, compared to an unstimulated control, from the same 4 patients as for the 10X transcriptomics. The TCR clonotypes found to be enriched by proliferation in the stimulated conditions compared to unstimulated were then matched to the same clonotype previously identified via 10X transcriptomics and overlayed on the same UMAP (Figure 7A).

**Figure 7.**
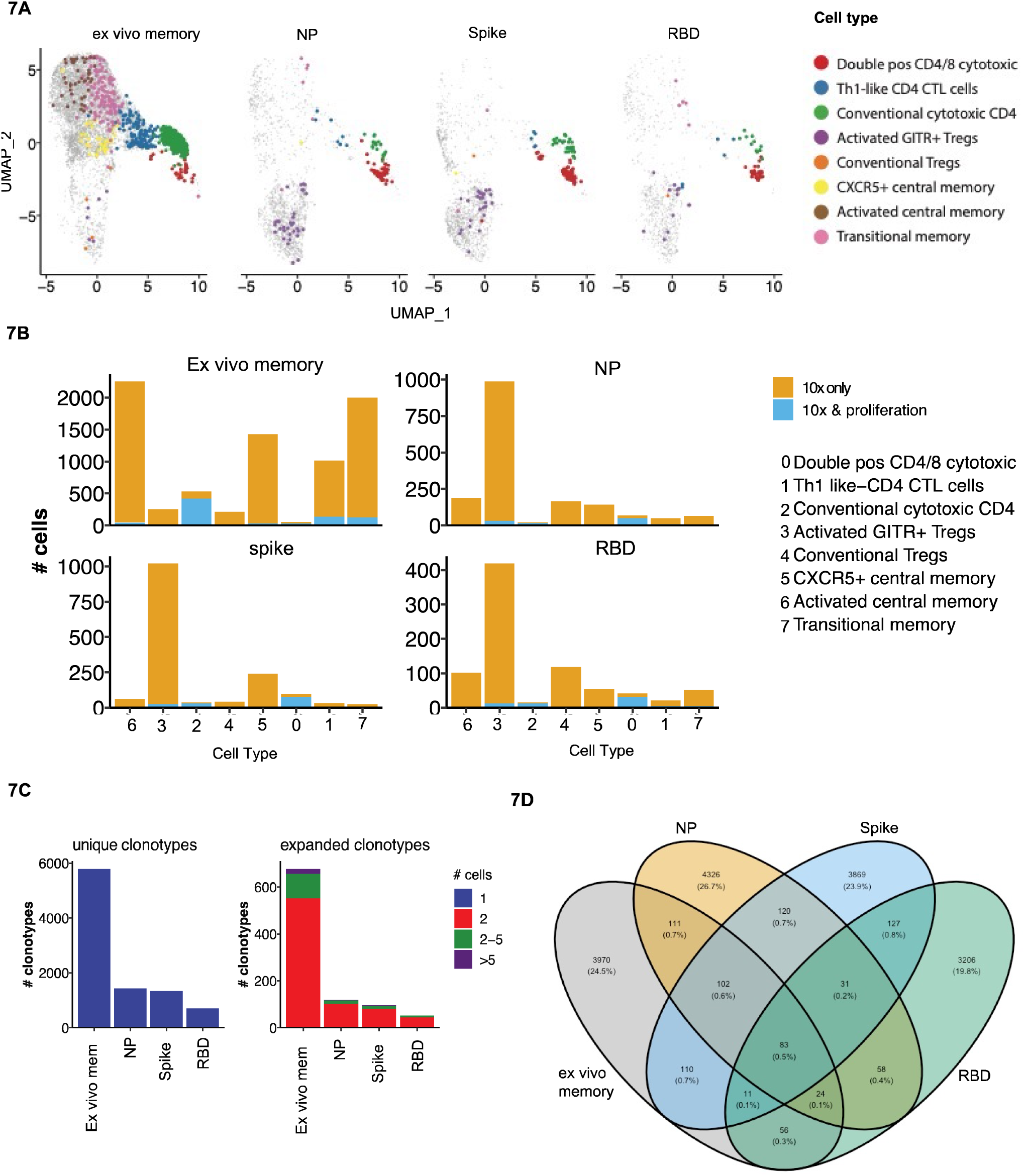
Proliferative clonotypes are enriched within Tregs and cytotoxic subsets. Bulk TCR sequencing of CD4s T cells from Ab high subjects that proliferated for 5 days following stimulation with SARS-CoV-2 peptide pools were matched with 10x single-cell TCRs to examine enriched clonotypes. **(A)** UMAP clonotypes that were enriched following proliferation and matched to 10x TCRs and split into 4 conditions: ex vivo memory, nucleoprotein (NP), Spike and RBD. **(B**) Proliferative CD4 clones were highly enriched with activated GITR+ Tregs and cytotoxic clusters. **(C)** Abundance of unique single clonotypes with few expanded clonotypes within stimulated conditions. **(D)** Minimal shared TCR clonotypes within stimulated conditions compared to ex vivo memory. Fishers exact test with Bonferroni’s corrected *p* values shown for log odd ratios.

10% of clonotypes were shared between the single cell RNAseq analysis and the proliferation assay TCR analysis. Enriched proliferative clonotypes identified in the SARS-CoV-2 antigen-stimulated cultures were most frequent in transcriptomic clusters comprising: (i) conventional Granzyme B+ CD4 CTL (cluster 2; Figure 7B), (ii) CD4/CD8 double positive CTL (cluster 0; Figure 7B); and (iii) relatively limited proliferation of activated GITR+ Tregs, compared to their identification in the OX40 assay 10x only analysis (cluster 3; Figure 7B). The majority of clonotypes identified through the single cell RNAseq analysis were unique singletons, while expanded clonotypes following proliferation consisted mostly of 2 cells with the same clonotype, as well as a small portion of clonotypes that were 2-5 cells (Figure 7C).

As expected, there was very little overlap of TCR clonotypes between the different antigen-specific responses, with <1% of clonotypes being shared (Figure 7D), consistent with high diversity within SARS-COV-2 reactive CD4 T cells. A total of 3, 599 unique SARS-CoV-2 clonotypes were identified in our study that have not been previously reported elsewhere.

### Increased RBD-specific CD4 T cells following vaccination

It was then important to assess whether vaccination would improve adaptive immune responses in the low antibody ADAPT convalescent subjects. Between 2 and 4 weeks following the vaccination second dose of either BNT162b2 or ChAdOx1, PBMC samples were collected from the previously infected participants, from either the high or low antibody patient groups, respectively. No longitudinal difference was observed between either spike or NP-specific CD4 T cell responses at D2 timepoint (2-4 weeks post-second dose) in both subject groups. However, in RBD-specific responses, in the original Ab low group, there was a 3.4-fold increase from the 8-month convalescent timepoint to the post-vaccination D2 timepoint (*p*<0.01; Figure 8A). No difference was observed in CD8 T cell response between pre or post vaccination timepoints (Figure 8B).

**Figure 8.**
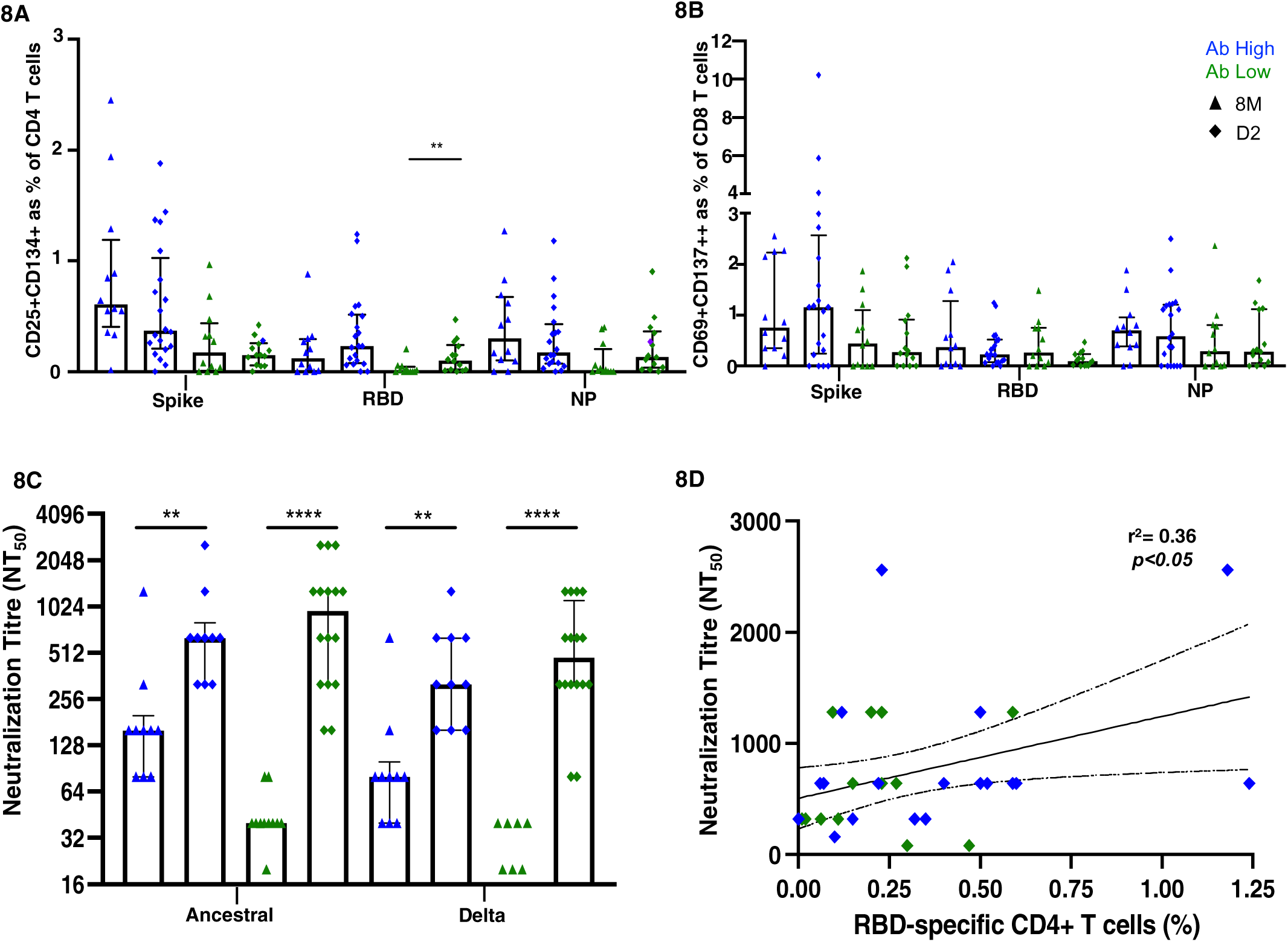
Vaccination induces RBD-specific CD4 T cells in Ab low subjects. **(A)** Significant increase in RBD-specific CD4+ T cells following vaccination in Ab low group. **(B)** No difference in CD8 response in both groups post-vaccination. **(C)** Increase in antibodies following two doses of SARS-CoV-2 vaccination that neutralized both ancestral and delta viruses in Ab low and Ab high groups. (**D)** Positive correlation between RBD- specific CD4+ T cells and neutralising antibody titres post second dose of vaccination. Data shown as medians with interquartile ranges. Two-tailed p values <0.05 were considered significant (*<0.05, **<0.01, ***<0.001. Mann- Whitney T tests were used for unpaired groups. Pearson’s rho was used for correlations and adjusted *p* values shown.

A sharp increase in neutralizing antibody titres towards the ancestral strain was observed in both Ab high and low groups after vaccination (Ab high 8M NT_50_=160, D2 NT_50_=640 [*p*<0.01]; Ab low 8M NT_50_=40, D2 NT_50_=960 [*p*<0.0001]). Neutralization of the B.1.617.2 delta strain was also increased (Ab high 8M NT_50_=80, D2 NT_50_=320 [*p*<0.01]; Ab low 8M NT_50_=0, D2 NT_50_=480 [*p*<0.0001]) (Figure 8C). As it was previously shown that low RBD-specific CD4 T cells response was associated with low neutralizing antibodies at 3- and 8-months post-infection, it was important to correlate these two parameters at the post-vaccination timepoint. A postive correlation (spearman’s rho=0.36, *p*<0.05) was observed between RBD-specific CD4 T cells in the OX40 assay and neutralization titre post-second dose (D2) timepoint (Figure 8D).

## DISCUSSION

This study has revealed that only those convalescent COVID-19 patients who had readily detectable RBD-specific CD4 T cell in vitro recall responses had significantly higher neutralizing antibody titres, whereas lower anti-spike antibody levels, especially neutralizing antibody titres, were associated with a lack of RBD-specific proliferative CD4 T cells . These results raise serious questions of the quality and quantity of any immediate anamnestic in vivo response in patients without these proliferative CD4 T cell responses, if re-exposed to SARS-CoV-2. In contrast, those patients with in vitro proliferative RBD-specific CD4 T cells would be predicted to mount an early vigorous in vivo immune response if re-exposed. Other groups using similar assays to our OX40 assay (*12–16*) have also found that the majority of recovered COVID-19 patients had detectable responses to pools of spike peptides, but with only a subset of patients having CD4 T cell responses to RBD epitopes. We have also previously shown that early CD4+ T cell responses can predict RBD-specific memory B cell frequencies at 1-year post-infection (*31*).

It is widely believed that neutralizing antibodies are the first line of defence against re-infection with the same viral pathogen (*4*). Our results agree with other studies that there is a very large variation in levels of such neutralizing antibodies in the serum of individuals recovered from COVID-19 (*28*); the reasons for this range of responses are unknown, but the quantity and quality of viral antigen-specific CD4 T cells are highly likely to be important (*5*). Furthermore, it is unknown what level of such antibodies are required for complete protection. However, with the current vaccines, and evidence from their Phase III trials, a correlation of higher neutralization titres and lower infection rates has been reported (*32*).

Our results directly correlate RBD-specific proliferative CD4 T cell responses in convalescent PBMC with anti-spike antibodies and neutralizing antibody titres. Furthermore, spike-specific CD4 T cells from recovered COVID-19 subjects with low antibody levels showed very little expansion in vitro, and in the smaller number of cells that did proliferate, they appeared to relatively highly express the Treg transcription factor Foxp3. Our CD25/OX40 assay was able to identify SARS-CoV-2-specific Tregs, using CD39 expression (*30*), which was confirmed at the transcriptional level with Foxp3 and GITR expression. Treg responses have been under-represented in other studies using other activation markers, namely CD40L (CD154) and 4-1BB (CD137) (*23, 33–35*), together with shorter incubations with antigens, that did not pick up these important regulatory cells. Further studies are needed to understand whether a preponderance of Tregs exerts a greater influence over a smaller number of spike-specific proliferation-capable CD4 T cells from the Ab low subjects.

When SARS-CoV-2 spike-, RBD- and NP-specific CD4 T cells from patients from the high Ab group were examined at the single cell transcriptomic level, heterogeneous profiles of CD4 T cell subsets were observed that included central memory cells and CD4 CTL, as well as regulatory T cell populations expressing GITR and Foxp3. Importantly, the specific CD4 and CD8 T cells from patients from the antibody low group showed a predominance of expression of Foxp3 and CTLA-4, respectively, associated with their limited proliferative responses. We have previously reported high CTLA-4 expression in HIV-specific CD4 T cells (*36*) which was inhibitory to proliferative responses in vitro (*37*), whereas vaccinia-specific CD4 T cells expressed less CTLA-4 and proliferated well, 21 days after inoculation (*7*).

T follicular helpher cells (Tfh) are vital in B cell maturation and immunoglobulin class switching within the germinal centre (*5*), Two studies have suggested that circulating Tfh (cTfh) from recovered COVID-19 patients include spike-specific CD4 T, but in one study RBD-specific cTfh were rare (*12*) while in the other, total spike-specific cTfh were only seen in 3/26 subjects (*38*), overall consistent with a large proportion of immunodominant epitopes for the spike-specific CD4 T cells being outside the RBD (*12, 39*). Interestingly, one study has reported an immunodominant CD4 epitope within RBD from convalescent PBMC, using in vitro proliferation in response to spike peptides, as an intitial step in a cloning procedure (*20*), but antibody levels were not reported.

We were able to characterize in detail the expanded RBD-specific CD4 T cells after in vitro proliferation and, surprisingly, we found no staining for the Tfh-defining transcription factor Bcl6 in the expanded cells from PBMC, despite using staining protocols that readily identified Bcl6+ Tfh and germinal centre B cells in other studies of lymph node (*40, 41*) and tonsil cells (*42*). In a previous study, we also identified *BCL6* at the single cell level in antigen-specific CD4 T cells in PBMC (*43*). Therefore, we postulated that spike-specific cTfh cells should be encompassed within the CXCR5+ and activated central memory subsets, observed from our single cell RNAseq data, but low transcript expression of *BCL6* did not give us a definitive answer. Nevertheless, future in vitro studies should examine whether memory CD4 T cells in PBMC, in particular cTfh, from recovered COVID-19 patients, can boost anti-spike antibodies by memory B cells in vitro, analogous to previous studies for other recall antigens (*44*). Instead, our analysis of proliferated RBD-specific CD4 T cells showed they were more likely to highly express T-bet, the Th1-defining transcription factor. Most studies have reported that IFN-γ is the most prominent cytokine produced by SARS-CoV-2 specific memory T cells in vitro in response to antigen (*12–15*), so expression of T-bet in expanded cells is consistent with a Th1 skewing of spike-specific effector CD4 T cells in PBMC.

We have previously shown that there are relatively high expression of cytotoxic lymphocyte markers in HIV-specific and CMV-specific CD4 T cells (*45–47*), as well as CD4 CTL found in other viral infections (reviewed recently in (*48*)). Very early following vaccinia inoculation, many vaccinia-specific CD4 T cells were also CTL, expressing mainly Granzyme K, at day 14, at the same time as the peak of activated CD4 T cells in blood (*8*), consistent with what was reported for a COVID-19 patient during the acute phase of the infection (*49*). We identified three transcriptomic clusters of cytotoxic CD4 T cells that were within the SARS-COV-2 reactive subsets by single cell RNAseq. This observation was consistent with our finding of in vitro expanded spike-specific CD4 T cells that expressed protein markers of cytotoxic T lymphocytes (CTL), including the granzymes A and B, and perforin, and cytotoxic granules (as defined by the antibody TIA-1, which recognizes the protein encoded by the gene *NKG7* (*45, 50*)). Highly enriched proliferative clonotypes were also shown to overlap with coventional CD4 CTL and CD4/8 double positive CD4 CTL by single cell RNAseq. It was also evident following bulk TCR-seq that proliferated clonotypes in respone to spike and RBD were enriched within CD4 CTL clusters. Effector CD4 T cells expressing cytotoxic granules have been identified in other single cell transcriptomic studies (*35, 51*), suggesting that CD4 CTLs may play an important role in eliminating SARS-CoV-2 infected cells.

Based on what is known about CD4 T cell help for B cell responses (*5*), it is highly likely that memory CD4 T cells in peripheral blood reflect a large CD4 T cell response in draining lymph nodes, including T follicular helper cells (Tfh), but also resulting in exit of antigen-specific effector and memory cells into the circulation. We have directly observed these highly activated, non-Tfh cells in lymph nodes in untreated HIV-1 infection (*40, 42*), but their clonal relationship to similarly elevated Tfh cells is still under study. From the current study, it isn’t clear at the molecular level why CD4 T cells with specificities to epitopes within RBD may generate better levels of anti-spike neutralizing antibodies. Theoretically, any spike-specific CD4 T cells should be able to help B cells that are able to take up spike protein via specificity for RBD. However, spike protein is very large and is designed to be cleaved during viral entry into target cells, involving multiple proteases (*52–54*), so it is possible that spike available for B cell recognition in germinal centres may also be cleaved to smaller protein fragments, only some of which link B cell and T cell epitopes around RBD. Clearly this needs further study with defined epitopes, along with the clonal relationships between germinal centre Tfh cells during acute infection and memory cells during convalescence. While early studies suggested that there can be intermolecular CD4 help for antibody responses in murine models of influenza infection (*55, 56*), in a vaccinia virus mouse model it was shown clearly that intramolecular CD4 help was required for antibodies to individual viral proteins from this large poxvirus (*57*). Most studies of SARS-CoV-2 specific T cells have used large peptide pools, whereas we have found most of our associations with recombinant proteins which may be an advantage because it requires the full antigen-processing and HLA Class II presentation pathway rather than extracellular saturation with a large number of exogenous peptides. Finally, it also has to be considered whether some RBD epitopes may be obscured by glycan residues that lock the RBD in a “down” position (*58*), since our recombinant proteins were made in human cell lines.

Possible reasons why some infected individuals have low adaptive immune responses could be due to (i) lower initial viral loads during first wave infections compared to later waves of variants (*59*); (ii) antigen presentation to CD4 T cells is dominated by epitopes in spike fragments outside of RBD (*39, 60*); and (iii) possibly having more inhibitory and/or exhausted B and T cells (*23*).

Importantly, however, it appears that RBD-specific CD4 T cells could be boosted by vaccination. Our results suggest that booster efficacy would be improved by concentrating on CD4 T cell epitopes in and around the RBD, which may be the optimal regimen for generation of neutralizing antibodies.

## MATERIALS AND METHODS

### ADAPT Cohort

The ADAPT study is a prospective cohort study of post-COVID-19 recovery established in April 2020 (*61*), with ongoing recruitment (147 participants with confirmed SARS-CoV-2 infection had been enrolled at the time of writing). The majority were recruited following testing in community-based clinics run by St Vincent’s Hospital Sydney, with some patients also enrolled with confirmed infection at external sites. Initial study follow-up was planned for 12 months post-COVID-19, and subsequently extended to 2 years. Extensive clinical data was systematically collected, including classification of disease severity, as previously described (*61*) and a prospective biorepository established, as previously described (*62*). Subjects classified with mild COVID-19 were those managed in the community with minor, largely upper respiratory tract viral symptoms, including pharyngitis, rhinorrhea, headache, and anosmia/ageusia. Subjects with moderate COVID-19 were managed in the community with fever/chills and one of the following organ-localizing symptoms, or at least two of the following organ-localizing symptoms: cough, hemoptysis, shortness of breath, chest pain, nausea/vomiting, diarrhea, altered consciousness/confusion. Subjects with severe COVID-19 were those who required inpatient care (wards or intensive care unit), as previously described (*61*). Laboratory testing for SARS-CoV-2 was performed using nucleic acid detection from respiratory specimens with the EasyScreen™ SARS-CoV- 2 Detection kit (Genetic Signatures, Sydney, Australia). Enrolment visits were performed at median 76 (IQR 64-93) days after initial infection (3-months) and 8-month assessments were performed at median 232 (IQR 226-253) days after initial infection. Serum and PBMCs were collected from ADAPT study participants following vaccination with either 2 doses of BNT162b2 (Pfizer) or ChAdOx1 (AstraZeneca).

The demographics of the ADAPT study participants in this report (first wave patients, recruited prior to October, 2020) are shown in Supplementary Table 1. Unexposed healthy adult donors (n=13; 45% male; median age 46) were recruited through St Vincent’s Hospital, were anti-spike antibody negative, and tested prior to vaccination.

### Ethics

The ADAPT study was approved by the St Vincent’s Hospital Human Research Ethics Committee (2020/ETH00964) and is a registered trial (ACTRN12620000554965). ADAPT-C sub study was approved by the same committee (2020/ETH01429). All data were stored using REDCap electronic data capture tools. Unexposed healthy adult donors were recruited through St Vincent’s Hospital which was approved by St Vincent’s Hospital Human Research Ethics Committee (HREC/13/SVH/145 and HREC/10/SVH/130). All participants gave written informed consent.

### Antigens

Recombinant SARS-CoV-2 RBD polypeptide was produced using DNA encoding a His-tagged 200 amino acid region, for residues 319 to 541 of SARS-CoV-2 S protein, as previously described (*63*), corresponding to the following amino acid sequence: RV QPTESIVRFP NITNLCPFGE VFNATRFASV YAWNRKRISN CVADYSVLYN SASFSTFKCY GVSPTKLNDL CFTNVYADSF VIRGDEVRQI APGQTGKIAD YNYKLPDDFT GCVIAWNSNN LDSKVGGNYN YLYRLFRKSN LKPFERDIST EIYQAGSTPC NGVEGFNCYF PLQSYGFQPT NGVGYQPYRV VVLSFELLHA PATVCGPKKS TNLVKNKCVN FG SHHHHHH, which was cloned into pCEP4 vector (Thermo Fisher) and expressed transiently in Expi293 cells (Thermo Fisher) using the Expifectamine transfection kit (Thermo Fisher). After 7 days of expression, the culture supernatant was filtered, dialysed against PBS and the protein purified using the His-tag and Talon resin (Thermo Fisher), as previously described (*63*).

Recombinant trimeric SARS-CoV-2 spike protein was expressed from a plasmid encoding the spike protein with C-terminal trimerization domain and His tag which was a gift from the Krammer lab (BEI Resources, NIAID, NIH). The plasmid was transfected into Expi293 cells and protein expressed for 3 days at 37°C, 5% CO2. The protein was purified using the His tag as for the RBD purification. The protein was further purified on a Superose 6 gel filtration column (GE Healthcare) using an AKTA Pure FPLC instrument (GE Healthcare) to isolate the trimeric protein and remove S2 pre-fusion protein, as previously described (*63*). Pools of 15-mer peptides corresponding to the sequence of SARS-CoV-2 spike protein were purchased from Genscript (Hong Kong) and are listed in Supplementary Table 2. His-tagged purified recombinant spike proteins from human coronavirus strains 229E, NL63 and OC43 were purchased from SinoBiologicals US (Wayne, PA, USA). Anti- CD3/anti-CD28/anti-CD2 polyclonal T cell activator was purchased from StemCell Technologies (Vancouver, Canada). Purified Influenza A (A/Sydney/5/97) was a gift from Alan Hampson, CSL, as previously described (*19*).

### CD25/OX40 assay

Peripheral blood mononuclear cells (PBMC) were isolated from EDTA anti-coagulated blood within 4 hr of venepuncture, as previously described (*8*). Antigen-specific CD4 T cells responding to recall antigens, simultaneously up-regulating CD25 and CD134 (OX40), were measured in cultures of 300,000 PBMC in 200 µl/well of a 96-well plate, in Iscove’s Modified Dulbecco’s Medium (IMDM; Thermofisher, Waltham, MA, USA) containing 10% human serum (kind gift, Dr Wayne Dyer, Australian Red Cross Lifeblood, Sydney, Australia), which were incubated for 44-48 hr, in a 5% CO_2_ humidified incubator, as previously described (*19*). Separate cultures were incubated with individual SARS-CoV-2 antigens as indicated. All experiments additionally included: (i) a culture medium only, negative control well; (ii) anti-CD3/anti-CD28/anti-CD2 T cell activator (1/200 dilution), polyclonal positive control well; and (iii) influenza virus (1/200 dilution), antigen positive control well. PBMC from the respective cultures were stained with CD3-PerCP-Cy5.5, CD4-FITC, CD25-APC, and CD134-PE (BD Biosciences, San Jose, CA, USA) and run on a 5-laser Fortessa X20 (BD Biosciences) as previously described (*64*). Antigen-specific CD4 T cells were gated and expressed as CD25+CD134+ % of CD4+ CD3+ T cells as previously described (*19*). Cultures were classified as positive for antigen-specific CD4 T cells if the CD25+CD134+ % of CD4+ CD3+ T cells was ≥ 0.2% (*65*).

### Cell Trace Violet (CTV) proliferation assays

PBMC were resuspended at a concentration of 10 x 10^6^/ml in PBS and incubated with Cell Trace Violet (CTV) dye (Thermofisher) at 5 µM for 20 min at RT, according to the manufacturer’s directions. Cells were washed once with 5x volume of IMDM/10% human serum and resuspended for cultures of 300,000 PBMC in 200 µl/well of a 96-well plate and incubated for 7 days in a 5% CO_2_ incubator. Different wells contained different antigens as indicated. All experiments also included: (i) culture medium only negative control well; (ii) anti-CD3/anti-CD28/anti-CD2 T cell activator (1/200 dilution) polyclonal positive control well; and (iii) influenza virus (1/200 dilution) antigen positive control well. After 7 days, cells from the respective cultures were stained with CD3-PerCP-Cy5.5, CD4-FITC, CD8-APC-H7 and CD25-APC (BD Biosciences), and analysed on a 5-laser Fortessa X20 (*64*) and antigen-specific CD4 T cells gated as CD3+CD4+CD25^high^CTV^dim^ as previously described (*66*). Cultures were classified as positive for antigen-specific CD4 T cells if the CD25^high^CTV^dim^ % of CD4+ CD3+ T cells was ≥ 1%.

### Intracellular analysis of transcription factors

Expanded CD25^high^CTV^dim^ antigen-specific CD4+ CD3+ T cells at the end of a 5 day incubation period were analysed for expression of intracellular markers including transcription factors, cytotoxic effector molecules and CTLA-4 using Transcription Buffer permeabilization reagents (BD Biosciences), according to the manufacturer’s directions. The monoclonal antibodies used were: Tbet-BV711 (BioLegend); RORgT-PE and CTLA-4-PECy5 (BD Bioscences); Eomes-PE-Cy7, Bcl6-PerCP-eFluor 701 and Foxp3-AF700 (eBiosciences, Thermofisher). Following intracellular staining, cells were resuspended in 1% paraformaldehyde/PBS and analysed on the 5-laser Fortessa X20.

### E*x vivo* phenotyping and combined CD4/CD8 T cell activation assay

Cryopreserved PBMCs were thawed using RPMI medium containing L-glutamine and 10% FCS (ThermoFisher Scientific, USA) supplemented with Penicillin/Streptomycin (Sigma-Aldrich, USW), and subsequently stained with antibodies binding to extracellular markers. Extracellular panel included: Live/Dead dye Near InfraRed, CXCR5 (MU5UBEE), CD38 (HIT2) (ThermoFisher Scientific, USA); CD3 (UCHT1), CD8 (HIL-72021), PD-1 (EH12.1), TIM-3 (TD3), CD27 (L128), CD45RA (HI100), IgD (IA6-2), CD25 (2A3), and CD19 (HIB19) (BioLegend, USA); CD4 (OKT4), CD127 (A019D5), HLA-DR (L234), GRP56 (191B8), CCR7 (G043H7) and CD57 (QA17A04) (BD Biosciences, USA).

FACS Perm Buffer II (BD Pharmingen) was used for intracellular staining of granzyme B (GB11, BD Biosciences). FACS staining of 48hr activated PBMCs was performed as described previously, but with the addition of CD137 (4B4-1) to the cultures at 24hrs. Final concentration of 10µg/mL of SARS-CoV-2 peptide pools (Genscript) were used, 1 µg/mL of Influvac tetra influenza vaccine (Mylan Health, Sydney, Australia) was used as a control antigen and staphylococcal enterotoxin B (SEB; 1 µg/ml) was used as a positive control (Thermo Fisher Scientific). In vitro activation mAb panel included: CD3 (UCHT1), CD4 (RPA-T4), CD8 (RPA-T8), CD39 (A1), CD69 (FN50) all BioLegend, CD25 (2A3), CD134 (L106)- BD Biosciences. Samples were acquired on the Aurora CS spectral flow cytometer (Cytek Biosciences, USA) using the Spectroflo software. Prior to each run, all samples were fixed in 0.5% paraformaldehyde. Data analysis was performed using FlowJo version 10.7.1 (TreeStar).

### Anti-Spike diagnostic serology

Antibodies to SARS-CoV-2 spike in serum samples from ADAPT subjects were measured using the LIAISON® SARS-CoV-2 S1/S2 IgG diagnostic assay (Diasorin, Saluggia, Italy), according to the manufacturer’s direction. This method quantitatively detects IgG anti-S1 and anti-S2 specific antibodies by indirect chemiluminescence immunoassay, using recombinant S1 and S2 antigens.

### Flow cytometry based IgG/IgM serology

The assay to detect patient serum antibodies against SARS-CoV-2 antigens using flow cytometry has been previously described in detail (*28*). Briefly, HEK293 cells were transfected to transiently express SARS-CoV-2 full-length Spike (Wuhan-1 D614), Membrane and Envelope proteins. Diluted patient serum was added to the cells, followed by adding AlexaFluor 647-conjugated anti-human IgG (H+L) (Thermo Fisher Scientific) or anti-human IgM (A21249, Thermo Fisher Scientific). The LSRII flow cytometer (BD Biosciences, USA) was used to acquire the cells and patients were confirmed SARS-CoV-2 antibody positive if, in at least two of three quality-controlled experiments, their median fluorescence intensity (ΔMFI LJ = LJ MFI transfected cells − MFI untransfected cells) was above the positive threshold (mean ΔMFI + 4SD of 24 pre-pandemic controls). Data were analysed using FlowJo 10.4.1 (TreeStar, USA), Excel (Microsoft, USA), and GraphPad Prism (GraphPad Software, USA).

### Live virus neutralization assay

HEK293T cells were transduced with lentiviral particles to stably express human ACE2 and TMPRSS2. Briefly, the lentiviral expression vectors pRRLsinPPT.CMV.GFP.WPRE (Follenzi et al., 2002) and pLVX-IRES-ZsGreen (Clontech) were used to clone the ORFs for hACE2 (Addgene#1786) and hTMPRSS2a (Addgene#53887, synthetic gene fragment;IDT), respectively. Lentiviral particles for transduction, to express the above proteins, were produced by co-transfecting the second generation lentiviral packaging constructs psPAX2 (courtesy of Dr Didier Trono through NIH AIDS repository) and VSVG plasmid pMD2.G (Addgene#2259) and the expression plasmids individually in HEK293T cells (Life Technologies) using polyethyleneimine, as previously described (*67*). To generate the HEK293/ACE2/TMPRSS2a cells, two successive rounds of lentiviral transductions were performed; the highly permissive clone, HekAT24 was identified by clonal selection and then used to carry out the SARS-CoV-2 neutralisation assay as previously described (*67*).

To perform the assay, HekAT24 cells were trypsinized and while in suspension stained with Hoechst- 33342 dye (5% v/v) (NucBlue, Invitrogen) and then seeded in a 384-well plate (Corning #CLS3985) at 16,000 cells per well in 40µL of DMEM-5% FCS. Patient plasma samples were mixed at two-fold dilutions with an equal volume of SARS-CoV-2 virus solution (4 × 10^3^ TCID_50_/ml). Following incubation at 37°C for 1 hour, 40µL were transferred in duplicate to the cells (final MOI = 0.05). The following variants of concern were included as viral variants: Beta (B.1.351), Gamma (P.1), Delta (B.1.617), as well as control virus from the same clade with matching ‘D614G’ background (B1.319). Following 24 hours of incubation, entire wells were imaged by high-content fluorescence microscopy and an automated image analysis software obtained the cell counts. The following formula was used to calculate the percentage of virus neutralisation: %N = (D-(1-Q)) × 100/D, where Q = nuclei count normalized to mock controls and D = 1-Q for average of infection controls as previously described (*67*). Neutralisation activity, NT_50_ was defined as the serum dilution that led to 50% neutralization of infection.

### Dimensional Reduction and Clustering Analysis

FCS3.0 files were compensated manually using acquisition-defined matrix as a guide, and gating strategy was based on unstained or endogenous controls. Live singlets were gated from proliferated CD25+CTV- CD4+, or CD8+, respectively, CD3+ T cells using FlowJo v.10.7.2, samples were decoded and statistical analysis between groups and unsupervised analysis was performed. For unsupervised analysis, the following FlowJo plugins were used: DownSample (v.3), UMAP (v.0.2), Phenograph (v.3.0) and ClusterExplorer (v.1.5.9) (all FlowJo LLC). Equal number of events for each condition were taken from each grouped sample by down sampling. The two new FCS files corresponding to Ab high and Ab low were then concatenated for dimensionality reduction analysis using UMAP. UMAP was conducted using the following parameters for proliferated CD4 T cells: T-bet, Eomes, Granzyme B, Foxp3, RORγt, and BCL-6; and for CD8+ T cells: T-bet, Eomes, Granzyme B and CTLA-4. The Phenograph plugin was then used to determine clusters of phenotypically related cells. The same markers as TriMap and parameters k = 152 and Run ID = auto was used for analysis. Finally, ClusterExplorer plugin was used to identify the phenotype of the clusters generated by Phenograph.

### Single cell RNA-seq analysis of antigen-specific T cells

PBMC from 4 Ab high donors were cultured for 48 hr with NP, spike and RBD peptide pools, respectively (Supplementary Table 2), as for the OX40 assays (see above). A total of 14,053 SARS-COV-2 specific CD25+OX40+ CD4 T cells from these cultures were purified using FACSAriaIII cell sorter (BD biosciences), as shown in Supplementary Fig 2A. Unstimulated purified CD45RO+ ex vivo memory CD4 T cells were used as a comparator subset (Supplementary Fig 2B). Purity of sorted populations were >99%. Populations were individually stained with Total-Seq C hashtags (BioLegend), and single cell libraries were generated using the 5’v2 Gene expression and immune profiling kit (10x Genomics). Subsequent cDNA and TCR libraries were generated according to manufacturer’s instructions. Generated libraries were sequenced on the NovaSeq S4 flow cell (Illumina) at Read 1 = 28, i7 index = 10, i5 index = 10 and Read 2 = 90 cycles according to manufacturer’s instructions.

### Transcriptomic analysis

#### Pre-processing of raw sequencing files

Single-cell sequencing data was aligned and quantified using Cell Ranger (10x Genomics) against the human reference genome (10x Genomics, July 7, 2020 release) with default parameters. Raw hashtag data was processed using CITE-Seq count algorithm (*68*). Cell conditions were demultiplexed using ‘HTODemux’ function implemented in Seurat (*68*). Filtering and quality control was performed using Seurat (*69*) on data containing 15,276 cells where 14,053 cells were retained satisfying thresholds of both <10% mitochondria content and number of genes between 300 and 5000. ‘SCTransform’ was used for normalization (*70*). Genes encoding BCR and TCR were removed to improve cell clustering. Principal Component Analysis (PCA) dimensional reduction was performed on variable genes identified by ‘VariableFeatures’ function and cell clusters were visualised using Uniform Manifold Approximation and Projection (UMAP) clustering implemented in Seurat (*69*).

#### Annotation of cell identities and Differential gene expression analysis

T cell sub-populations were manually annotated based on UMAP clustering and markers defined by ‘FindAllMarkers’ function in Seurat.

Raw counts from defined cell populations were normalised using scran / scater (*71, 72*) and differential gene expression analysis was performed using Limma voom (*73*).

### TCR analysis of bulk proliferated cells

RNA was extracted from bulk proliferated cultures using RNAeasy micro kit (Qiagen) as per manufacturer’s instructions. TCR high throughput RNA sequencing methods have been previously published in detail (*74*). Briefly, reverse transcription of RNA was performed using a modified SmartSeq2 protocol that incorporated a 10bp universal molecular identifiers (UMI) into cDNA molecules. The first round of PCR was performed using primers against adapter sequences incorporated during cDNA synthesis with 1x KAPA HiFI HotStart ReadyMix and 8.3 mM Fwd and Rev primer with the following conditions: 98°C for 3 min; [98°C for 20 s, 67°C for 15 s, 72°C for 6 min] x 10 cycles; 72°C for 5 min. Purified PCR products were used in a second PCR targeting the TCRβ chain under the following conditions: 98°C for 45 s; [98°C for 15 s, 60°C for 30 s, 72°C for 30 s] x 30 cycles; 72°C for 1 min. Purified PCR products were then barcoded using the Nextera Index kit to enable pooling of multiple samples and sequenced on an Illumina MiSeq at 300bp paired-end reads to a depth of ∼0.5 million read pairs per sample. Primer sequences were as previously described (*74*).

### Analysis of single cell and bulk TCR datasets

Bulk TCRB datasets for the proliferation assay were processed with the Presto package (version 0.7.0 2021.10.28) (*75*). Reads were filtered for a minimum quality score of 20 using FilterSeq. R2s were trimmed of the TRC primer using MaskPrimers requiring exact primer matches. MaskPrimers was also used to extract the 10 nucleotide UMIs from R1 and to trim to TSO sequences. Trimmed R1 and R2 were paired with PairSeq and consensus UMIs were defined with BuildConsensus. R1 and R2 were then merged with AssemblePairs and the dataset was dereplicated to unique sequences and converted from fastq to fastq with CollapseSeq. Sequences with ambiguous bases (n) were discarded. Dereplicated fasta datasets were aligned against the IMGT human TCR reference directory [https://www.imgt.org/vquest/refseqh.html, downloaded 16-Jan-2020] using IgBLAST (*76*) via AssignGenes to generated AIRR formatted output.

To ensure consistent gene calling, TCR contigs from 10x’s cellranger vdj were re-aligned to the IMGT human TCR reference set with AssignGenes from the Presto package. Change-o databases were generated with MakeDb and subset to TCRA and TCRB with ParseDb from the Change-o package (version 1.2.0 2021.10.29) (*77*).

TCR clonotypes were defined using the TCRV, TCRJ and CDR3 AA sequence. Clonotype information from 10x VDJ was integrated with the 10x scRNA-seq within the Seurat package (version 4.1.0) (*78*) in R [R Core Team (2020). R: A language and environment for statistical computing. R Foundation for Statistical Computing, Vienna, Austria. URL https://www.R-project.org/] using RStudio [RStudio Team (2021). RStudio: Integrated Development Environment for R. RStudio, PBC, Boston, MA URL http://www.rstudio.com/] based on shared cell barcodes. For the proliferation assay bulk sequencing, enrichment was calculated by comparing clonotype UMI counts in stimulation conditions versus baseline/unstimulated with a pseudocount of 1 for clonotypes that were absent from the baseline sampling. Proliferation assay data was merged with the 10x data based on shared TCRB clonotypes.

TCR repertoires were explored using the tidyverse package (*79*) to aggregate and summarise data. Diversity was calculated as Shannon’s Entropy (*80*) as implemented by the entropart package (*81*). Repertoire features were visualised with ggven [https://CRAN.R-project.org/package=ggvenn], circos (*82*), ggpubr [https://CRAN.R-project.org/package=ggpubr] and ggsci [https://CRAN.R-project.org/package=ggsci].

To annotate previously reported SARS-CoV-2 specific T cells, TCRBs reported to bind SARS-CoV-2 epitopes were obtained from two public resources; immuneCODE MIRA (release 002.2) (*83*) and VDJdb (v2021-09-05) (*84*). TCRB clonotype labels were reformatted for consistency and clonotypes were annotated based on shared clonotype labels between the databases and dataset.

### Statistical Analysis

All column graphs are presented as medians with inter-quartile ranges. Mann-Whitney non-parametric test was used to compare unpaired groups and Pearsons’ correlation was used to analyse statistical relationships between continuous variables, employing Prism 9.0 software (GraphPad, La Jolla, CA, USA). RStudio version 1.2.1335 was used to generate PCA graphs. *p* values <0.05 were considered significant (*<0.05, **<0.01, ***<0.001, and ****<0.0001).

### Data availability

Single-cell RNA transcriptomic and TCR sequences including codes and raw data has been deposited to NCBI GEO database, accession number GSE198281. Novel SARS-CoV-2 specific TCR sequences described in our study will be deposited into VdJdB database.

## Data Availability

All data produced in the present study are available upon reasonable request to the authors

## SUPPLEMENTARY DATA

Supplementary Table 1 - ADAPT study patient characteristics. Age, gender, ethnicity, and severity at month 3 and 8 post-infection.

Supplementary Table 2 - Peptide pools: SN (spike non-RBD), SR (spike RBD), NP (Nucleocapsid protein).

Supplementary Figure 1 - Cell trace violet (CTV) 7-day proliferation assay.

Supplementary Figure 2 - CD39+ antigen-specific Treg responses.

Supplementary Figure 3 - Cell sorting for single-cell transcriptomics.

Supplementary Figure 4 - Diversity of SARS-CoV-2 specific TCR clonotypes.

## ACKNOWLEDGMENTS

The authors thank Bertha Fsadni, Sri Meka, Julie Jurczyluk and Kate Merlin at the St Vincent’s Institute for Applied Medical Research-Clinical Trials Unit for their expertise in specimen processing and bio-banking. We thank Dr Emma Johansson Beves for assistance on the Cytek Aurora.

## Funding

St Vincent’s Clinic Foundation Grant (GVM, GJD, DD) Medical Research Future Fund Grant (ST, FB-T, TGP, JZ)

National Health and Medical Research Council (NHMRC) Program Grant ID1149990 (ADK) NHMRC Fellowship ID1155678 (TGP)

UNSW Cellular Genomics Futures Institute Seed Funding (WHK)

Mrs. Janice Gibson and the Ernest Heine Family Foundation (PIC and TGP) Garvan Institute COVID Catalytic Grant (TGP)

UNSW COVID-19 Rapid Response Research Initiative (TGP)

## Author contributions

Protocol design and clinical management: GVM, GJD, DD

Experimental design and procedures: CP, WHK, KLJ, VK, AH, AAg, AAk, VM, AOS, RR, PS, MLK, HL, TD, MS, CMLM, MS, FB-T, ST, TGP, ADK, JZ

Visualization: CP, WHK, KLJ, JZ

Funding acquisition: GVM, GJD, DD, DC, ST, FB-T, TGP, PC, ADK, JZ

Supervision: DC, ST, PC, GVM, PIC, TGP, ADK

Writing – original draft: CP, WHK, KLJ, TGP, JZ

Writing – review & editing: CP, WHK, KLJ, CMLM, ST, AAg, TGP, ADK, JZ

## Supplementary Figures

**Supplementary Table 1.**
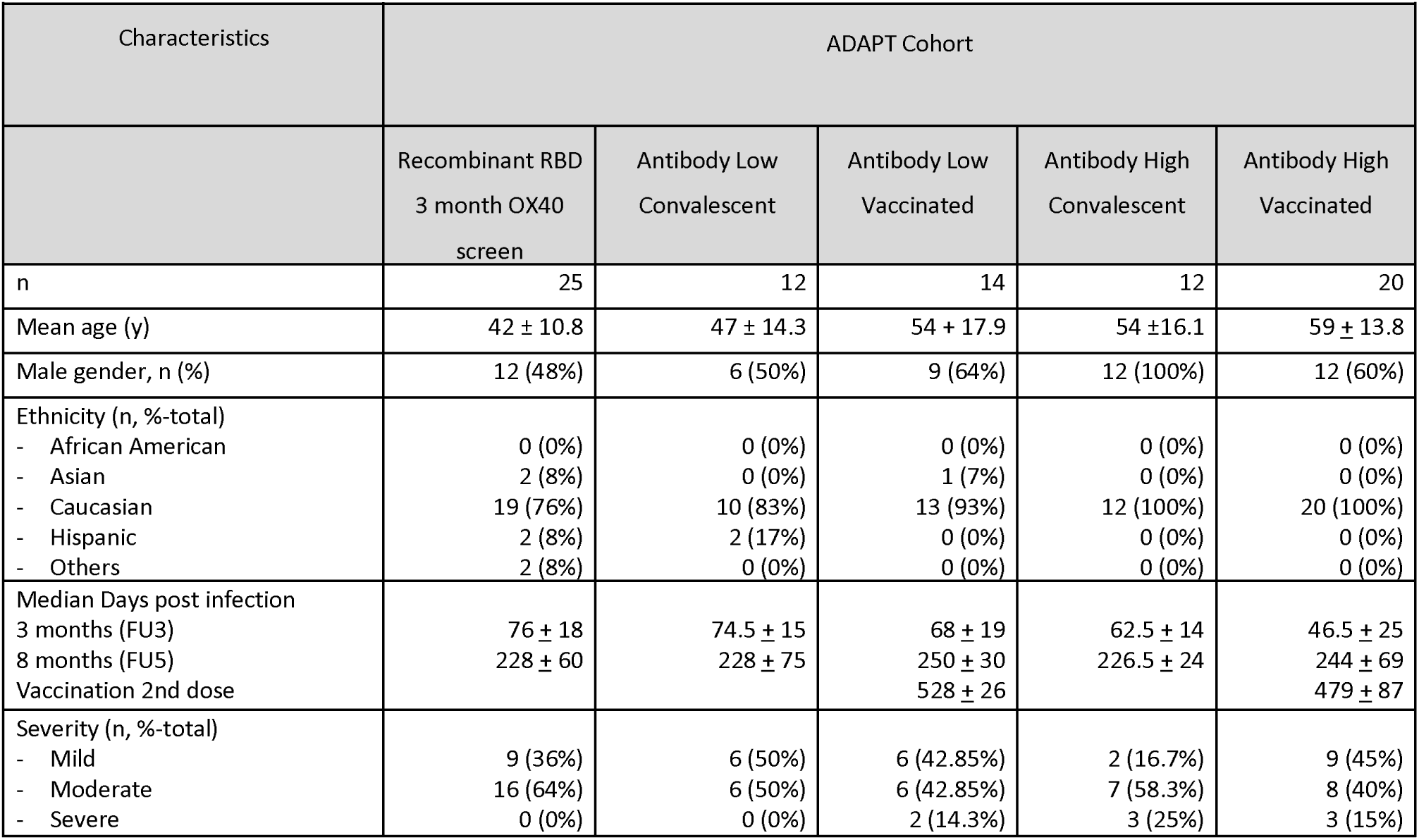
*ADAPT study patient characteristics.* Age, gender, ethnicity, severity, and medi neutralisation titre at month 3.

| Characteristics | ADAPT Cohort |  |  |  |  |
| --- | --- | --- | --- | --- | --- |
|  | Recombinant RBD<br>3 month OX40<br>screen | Antibody Low<br>Convalescent | Antibody Low<br>Vaccinated | Antibody High<br>Convalescent | Antibody High<br>Vaccinated |
| n | 25 | 12 | 14 | 12 | 20 |
| Mean age (y) | 42 ± 10.8 | 47 ± 14.3 | 54 ± 17.9 | 54 ± 16.1 | 59 ± 13.8 |
| Male gender, n (%) | 12 (48%) | 6 (50%) | 9 (64%) | 12 (100%) | 12 (60%) |
| Ethnicity (n, %-total) |  |  |  |  |  |
| - African American | 0 (0%) | 0 (0%) | 0 (0%) | 0 (0%) | 0 (0%) |
| - Asian | 2 (8%) | 0 (0%) | 1 (7%) | 0 (0%) | 0 (0%) |
| - Caucasian | 19 (76%) | 10 (83%) | 13 (93%) | 12 (100%) | 20 (100%) |
| - Hispanic | 2 (8%) | 2 (17%) | 0 (0%) | 0 (0%) | 0 (0%) |
| - Others | 2 (8%) | 0 (0%) | 0 (0%) | 0 (0%) | 0 (0%) |
| Median Days post infection |  |  |  |  |  |
| 3 months (FU3) | 76 ± 18 | 74.5 ± 15 | 68 ± 19 | 62.5 ± 14 | 46.5 ± 25 |
| 8 months (FU5) | 228 ± 60 | 228 ± 75 | 250 ± 30 | 226.5 ± 24 | 244 ± 69 |
| Vaccination 2nd dose |  |  | 528 ± 26 |  | 479 ± 87 |
| Severity (n, %-total) |  |  |  |  |  |
| - Mild | 9 (36%) | 6 (50%) | 6 (42.85%) | 2 (16.7%) | 9 (45%) |
| - Moderate | 16 (64%) | 6 (50%) | 6 (42.85%) | 7 (58.3%) | 8 (40%) |
| - Severe | 0 (0%) | 0 (0%) | 2 (14.3%) | 3 (25%) | 3 (15%) |

**Supplementary Table 2.**
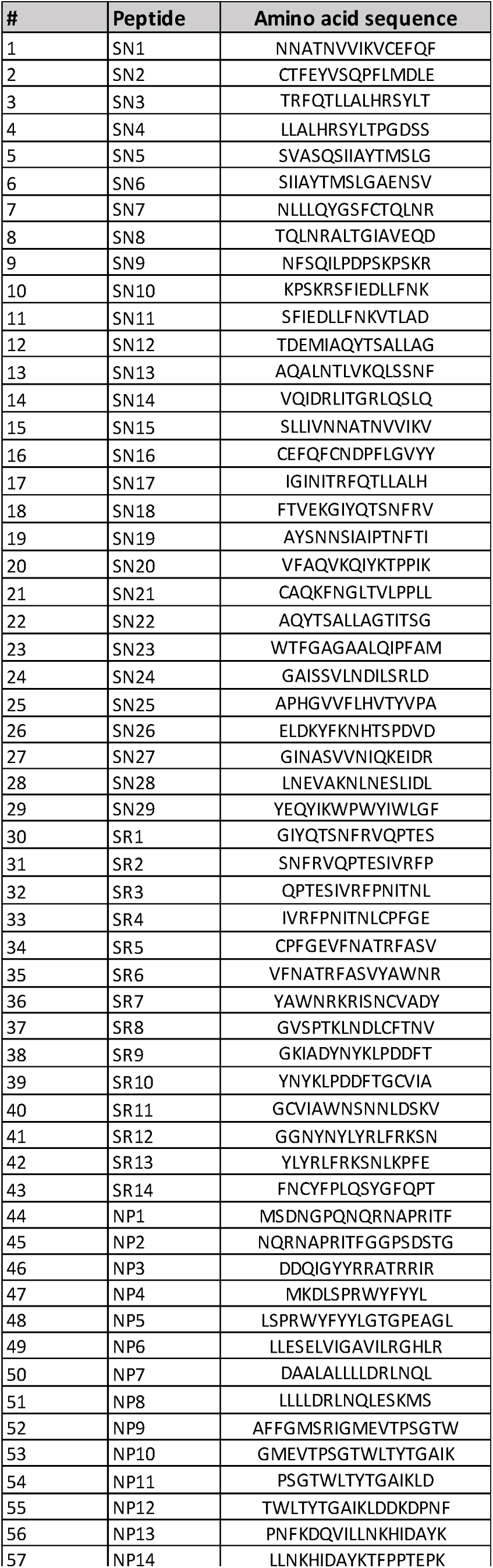
Peptides. SN (spike), SR (RBD), NP (Nucleocapsid protein).

**Supplementary Figure 1.**
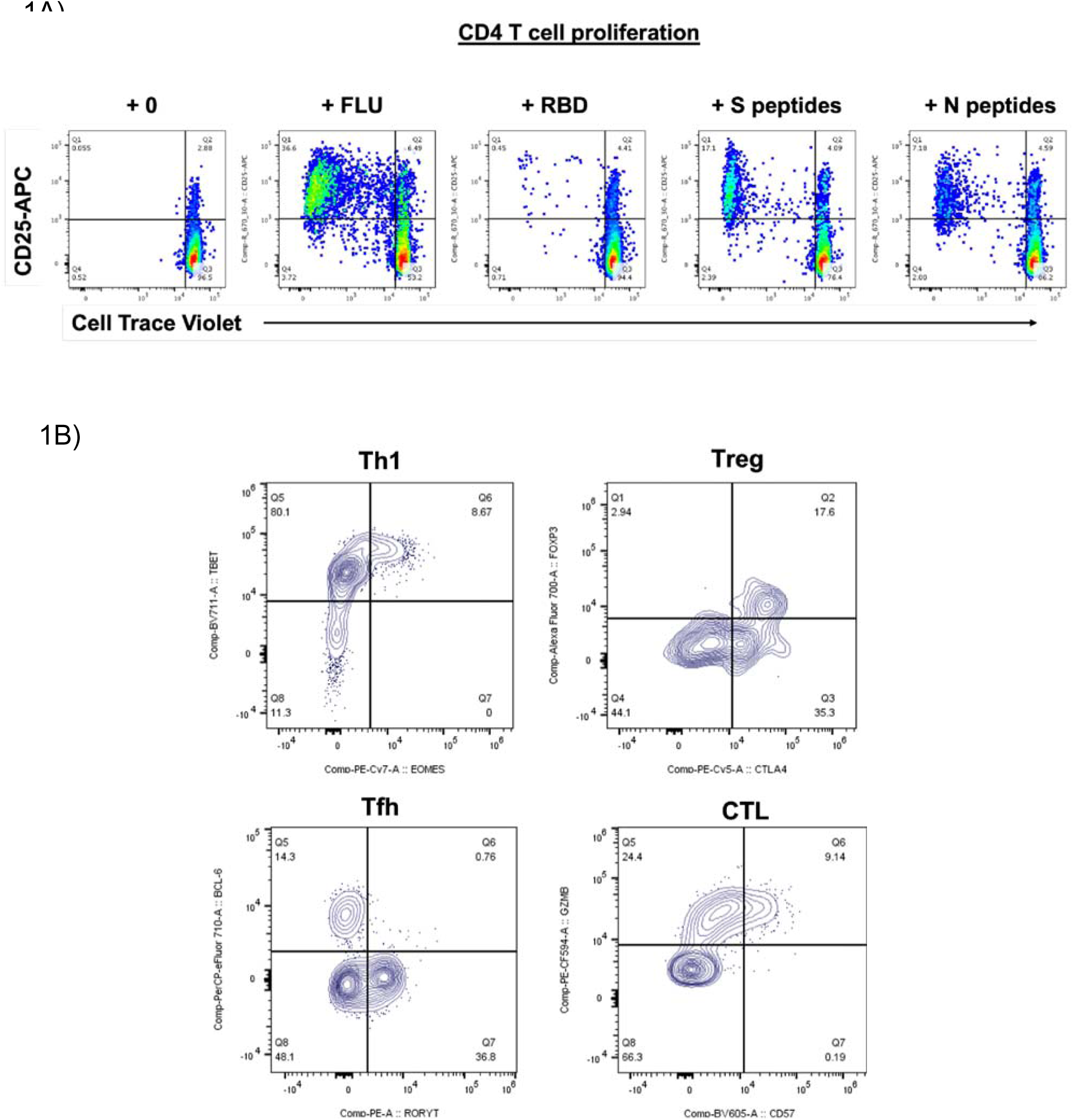
**T cell proliferation and phenotyping.** (A) Proliferated cells at day 7 in response to antigen stimulation in Q1 CD25+CellTraceViolet- (B) Gating strategy of proliferated T cells using transcription factors and activation markers.

**Supplementary Figure 2.**
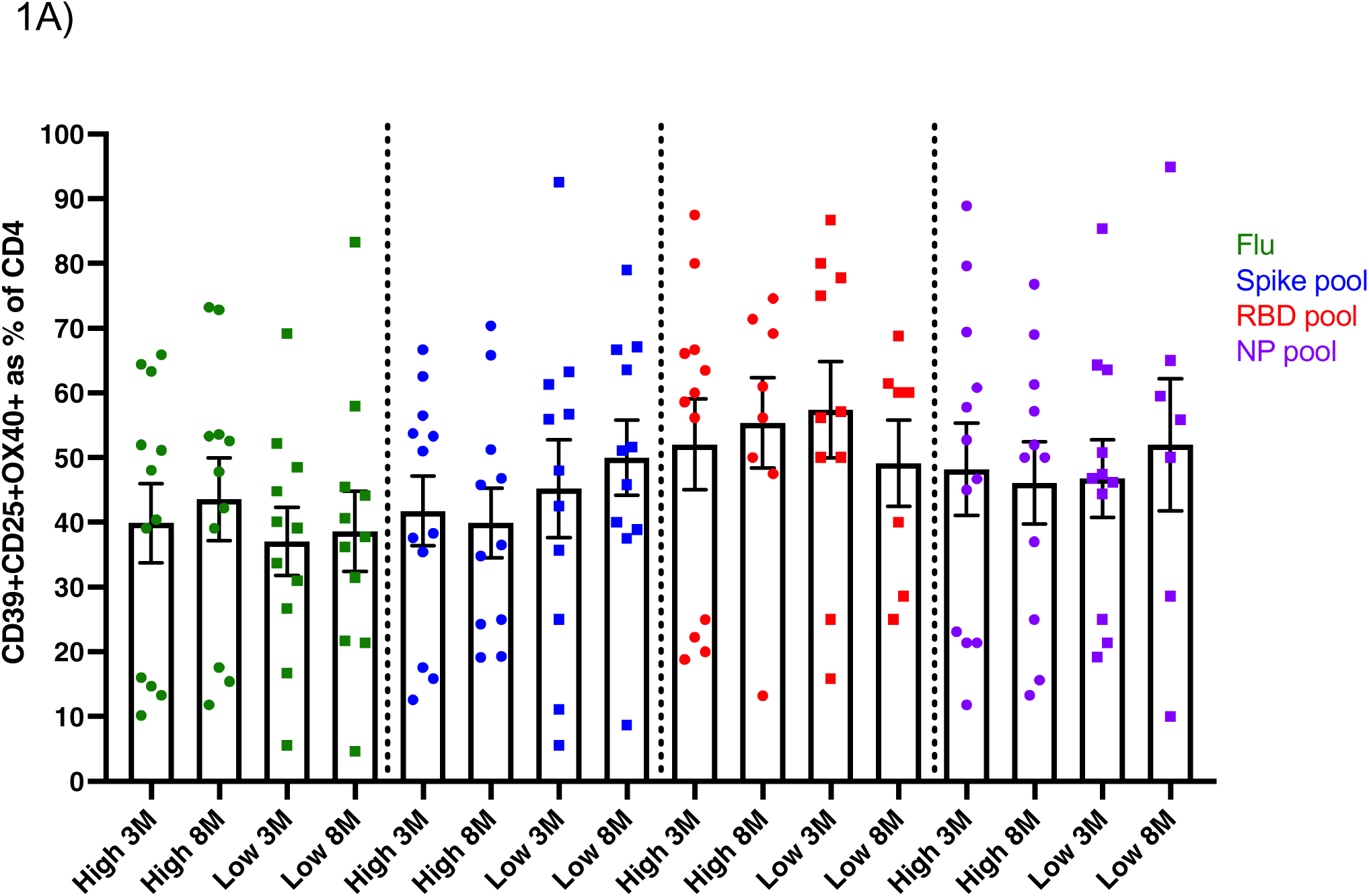
CD39+ antigen-specific Treg responses. **(A)** No difference was observed in CD39+ antigen-specific Tregs (CD25+CD134+CD4+) following stimulated with flu or SARS-CoV-2 peptide pools.

**Supplementary Figure 3.**
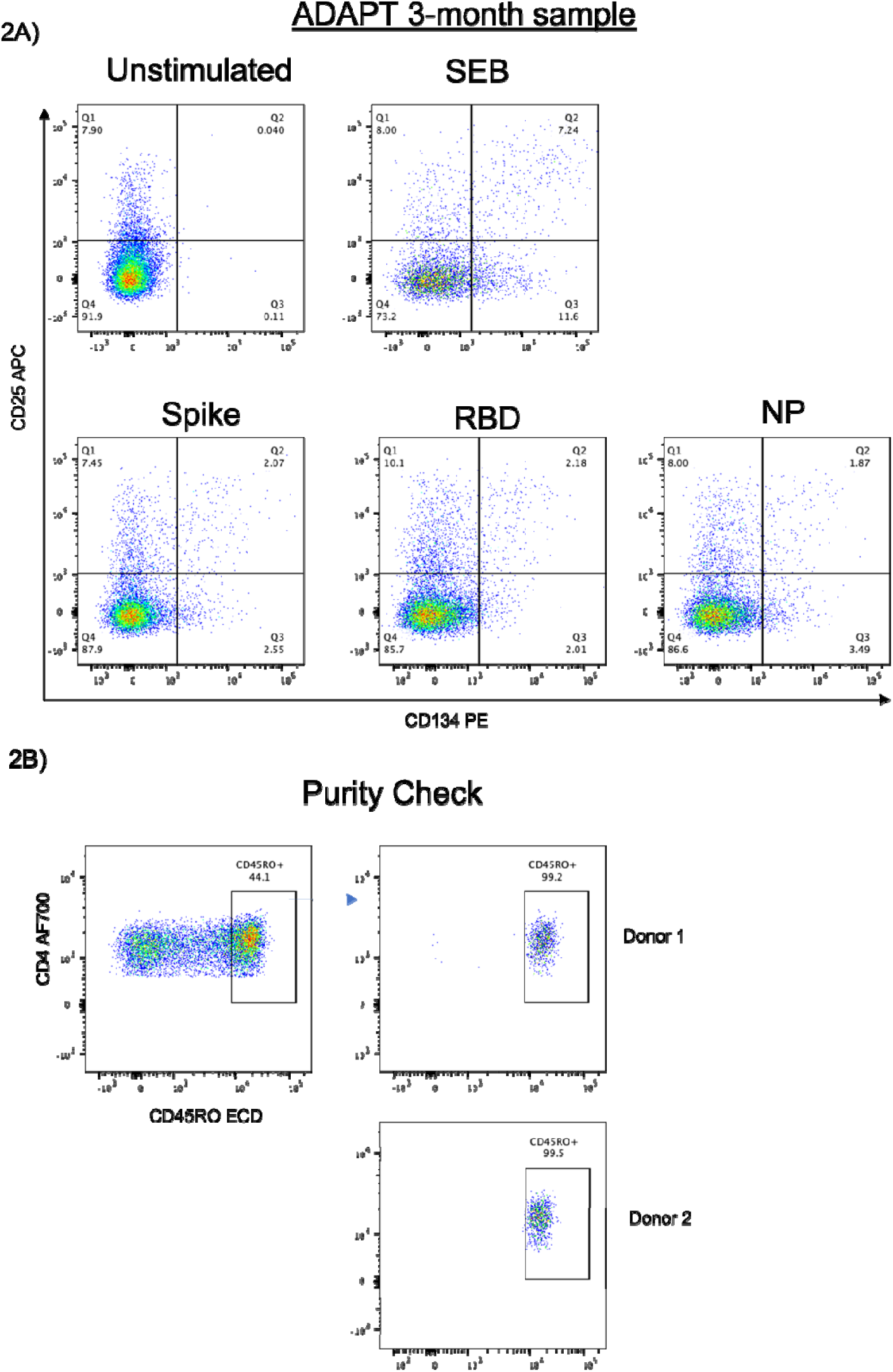
Cell sorting for single cell transcriptomics. (A) Representative dot of 1 partcipant shown followlng 48hr stimulatlon with SAR-CoV-2 peptide poole, with SEB as a positive control. Antigen-specific CD4+ T cells were bulk sorted from Q2 (CD25+CD134+) on the BD Aria Ill. (B) CD46RO+ ex vivo (unattmulated) memory CD4+ T cells were also bulk sorted far 10X sequencing. A purity of >99% was achieved for all sample.

**Supplementary Figure 4.**
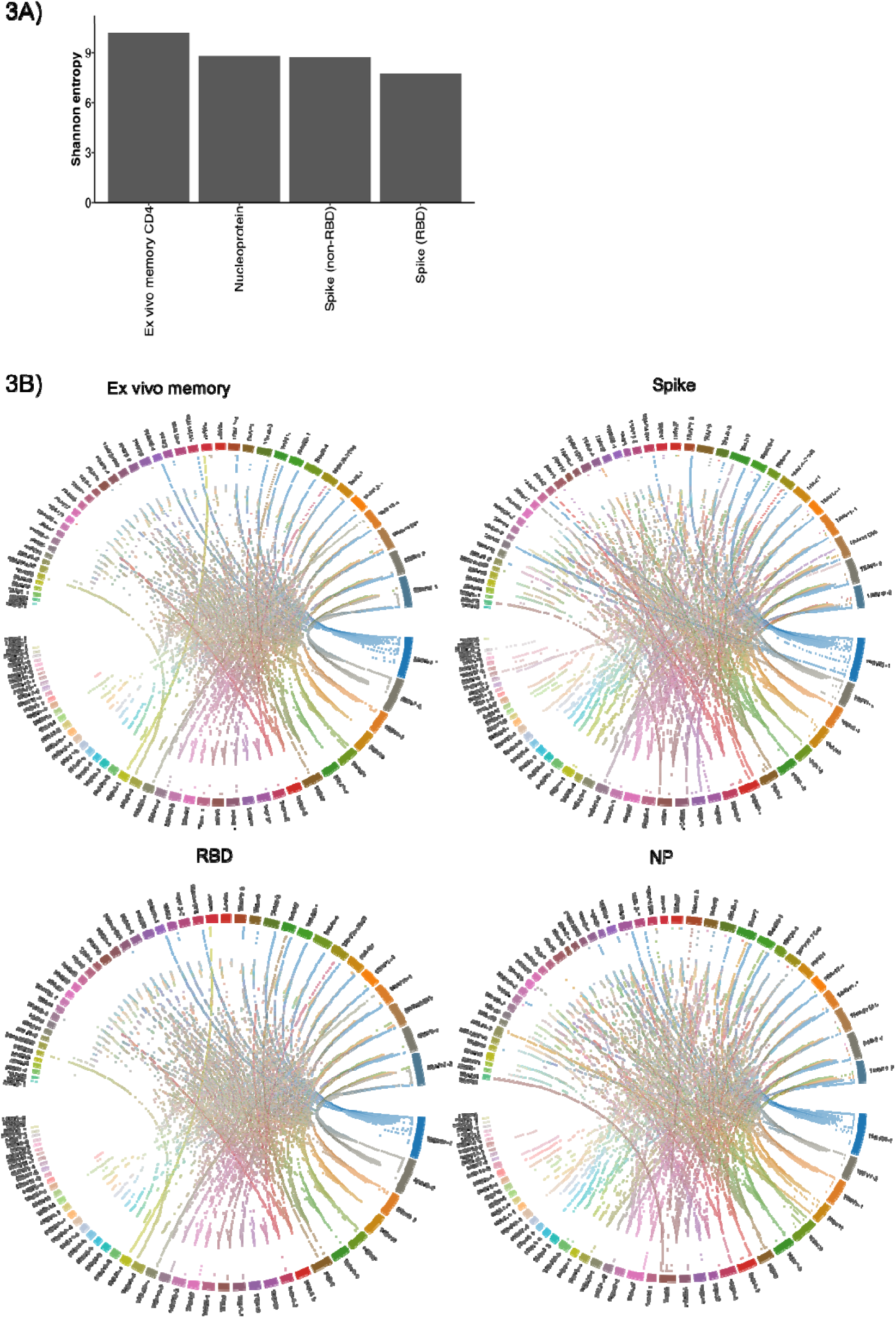
Diversity of SARS-CoV-2 specific TCR clonotypes. (A) Slightly lower Shannon’s entropy scores for spike, RBD end NP compared to ex vivo memory. (B) Circos plot showing high diversity of TCRαβ usage in all 4 conditions.

